# Systematic evaluation of semantic similarity across language models for schizophrenia spectrum disorders

**DOI:** 10.64898/2026.10.23.26366346

**Authors:** Amir H. Nikzad, Yan Cong, Ryan Partlan, Jiefei Li, Sunghye Cho, Sunny X. Tang

**Affiliations:** Northwell, New Hyde Park, NY; Department of Psychiatry, Zucker School of Medicine at Hofstra/Northwell, Hempstead, NY; Feinstein Institutes for Medical Research, Northwell Health, Manhasset, NY; School of Languages and Cultures, Purdue University, West Lafayette, IN; Department of Communication Sciences and Disorders, University of Cincinnati, Cincinnati

**Keywords:** Large Language Models, Psychosis, Natural Language Processing, Computational Psychiatry

## Abstract

**Background:** Semantic similarity features derived from language embeddings have been proposed as markers of schizophrenia spectrum disorders (SSD), but prior studies have reported inconsistent directions of effect and limited evidence regarding their relationship to specific dimensions of thought and language disturbance. We systematically evaluated how language model architecture and preprocessing strategy influence semantic similarity signals in SSD.

**Methods:** Speech samples from 228 participants (152 SSD, 76 healthy volunteers) were analyzed using six embedding models: GloVe, LSA, Word2Vec, RoBERTa, GPT-2, and Llama. Semantic similarity features were computed across three preprocessing levels (verbatim, disfluency-removed, and disfluency-removed with stopword filtering) using k-interword, moving-window, and sentence-to-sentence comparisons, summarized with various statistical descriptors. Features were compared between SSD and healthy volunteers and examined for associations with dimensions of thought and language disturbance, including inefficiency, incoherence, and impaired expressivity.

**Results:** SSD was predominantly associated with *increased* semantic similarity across models and comparison strategies. Model performance varied by preprocessing. RoBERTa performed best on minimally processed transcripts, yielding the greatest number of diagnosis-associated features, whereas GPT-2 achieved the largest diagnostic effect size after maximal preprocessing. GPT-2 showed the strongest signal for inefficiency and incoherence, while RoBERTa yielded the greatest number of associations with impaired expressivity.

**Conclusions:** SSD diagnosis and its dimensions of thought and language abnormalities are characterized primarily by greater semantic proximity in open-ended speech. Optimal preprocessing depends on model architecture and the clinical dimension of interest, highlighting the importance of model-and phenotype-specific analytic choices in computational studies of thought and language disturbance in SSD.

## 1. Introduction

Schizophrenia spectrum disorders (SSD) are marked by alterations in underlying meaning-making processes manifested in speech (1). Phenomenologically, alterations are often described along two dimensions: 1. Disorganization, in which the global or local structure of meaning is disrupted, leading to incoherent contents or tangential transitions (2,3); 2. Impoverishment, in which the effective semantic space is constricted, due to reduced lexical variety and repetitive or perseverative content (4). Increasingly, emerging natural language processing (NLP) methods have been implemented to objectively quantify and track symptom severity including language disturbances in SSD (5–8).

A leading NLP approach examines distances in meaning between linguistic units by leveraging semantic embeddings, which numerically represent the meaning content of linguistic units such as words and sentences and enable their comparison across a speech sample (9). Embeddings are extracted from language models which encode each linguistic unit as a vector in a multidimensional semantic space, allowing semantic relationships to be quantified geometrically, for example through cosine similarity.

Language models can be broadly divided into two categories: static vs. contextual. The first comprises static, or non-contextual, models, which assign each linguistic unit within a pre-determined dictionary a fixed representation independent of its surrounding context, such as Global Vectors for Word Representation (GloVe) (10), Latent Semantic Analysis (LSA) (11), and Word to Vector (Word2Vec) (12). The second type of language models comprises contextual models, which generate representations that depend on the linguistic environment of each unit, incorporating either preceding words only (unidirectional models, e.g., Generative Pre-trained Transformer 2, GPT-2 (13), and Large Language Model Meta AI, Llama (14)) or both preceding and following words (bidirectional models, e.g., Robustly Optimized Bidirectional Encoder Representations from Transformers Pretraining Approach, RoBERTa (15).

Numerical semantic representations extracted from language models could be then compared using variety of comparison strategies each generating a semantic similarity metric. Comparisons may be made at different levels of language organization for example between words (word-to-word similarity) or sentences (sentence-to-sentence similarity). Similarity may be captured between linguistic units at various intervals (e.g., comparing adjacent words or sentences, or those occurring at certain distance from each other), or within a block of word sequences (e.g., extracting similarities within a moving window of certain number of words). Finally, each semantic similarity metric may be described using different statistical approaches such as mean, median, minimum, maximum, etc., of semantic similarities (9).

Alteration in semantic similarity features has been associated with psychotic disorders including SSD. Earlier studies conceptualized *decreased* semantic similarity as a proxy for loosening of association, as occurs in disorganization phenotypes (16). Elvevåg et al. reported decreased semantic similarity in one-word associations, animal fluency task, and comparisons between participants’ responses and original questions in open-ended interviews, hence, coining the term *coherence* for cosine semantic similarity metrics (17). Similarly, decreased semantic similarity was observed in psychosis between adjacent sentences (sentence-to-sentence) and with respect to an artificial stimulus description (sentence-to-reference), ostensibly capturing incoherence and tangentiality (18). Other studies have reported reduced semantic similarity during animal fluency in patients with SSD (19), particularly among those exhibiting phenotypes of disorganization such as derailment or tangentiality (20). This pattern, however, was not replicated in a later study conducted by our team (21). Finally, decreased semantic similarity was predictive of conversion to psychosis in longitudinal clinical high risk (CHR) cohorts (22,23).

An opposing line of research has supported the inverse pattern, linking *increased* semantic similarity to psychosis (24). In this vein, increased mean semantic similarity, including higher average similarity across all word pairs and within moving windows of ten words during picture-description, was observed in first-episode psychosis compared to healthy controls (25). A subsequent study, reported higher word-to-word semantic similarity in picture description in psychosis despite no significant difference in word diversity or content specificity to stimuli, suggesting more constrained word selection from tightly related semantic clusters (26). With further replications of increased semantic similarity in psychosis (27,28), this phenomenon was interpreted as the reflection of narrowing of lexical-conceptual semantic space (29,30), challenging the earlier notion of semantic similarity as primarily an index of coherence (31).

Going beyond diagnostic differences, the relationship between semantic similarity and the dimensions of thought and language disturbance is not well understood. Despite conceptual elegance of interpreting decreased semantic similarity as a measure of decreased coherence of thought and language (17–19), there is stronger evidence pointing instead to a relationship between greater semantic similarity and impoverishment. Palominos and colleagues reported an association between greater alogia based on the Scale for the Assessment of Negative Symptoms (SANS) and word-to-word average and maximum semantic similarity using BERT model and minimum semantic similarity using FastText as well as negative correlation with maximum semantic similarity based on FastText (30). No word-to-word semantic similarity metric correlated with positive formal thought disorder based on the Scale for the Assessment of Positive Symptoms (SAPS). This result was consistent with the hypothesis that impoverishment involves narrowing of the effective semantic space as measured by contextual and non-contextual models (32). Other studies investigated indirect relationships between semantic similarity measures and formal thought disorder per their contribution to predictive models. Delal et al. found that semantic similarity was strongly predictive of impoverishment per Thought and Language Index (TLI) in relapse prediction Bayesian models, without finding such relationship with TLI disorganization (24). In a recent study by Ciampelli et al., a semantic similarity factor score showed poor performance in item-level symptom severity prediction: in the Dutch sample, semantic similarity factor failed to be significant across all mixed-effect models to predict The Positive and Negative Syndrome Scale (PANSS) items including conceptual disorganization. Similarly, word and sentence similarity factors showed no predictive signal for conceptual disorganization per Brief Psychiatric Rating Scale (BPRS) in the English sample (33).

Model selection, preprocessing approach, and feature curation introduce additional complexity in utilizing semantic similarity features for the detection of SSD. Language models have different intrinsic architecture that affects semantic representations obtained from them. Tokenization and preprocessing decisions may preserve, enhance or remove clinically relevant signals. Comparison strategies, as described above, can also influence the direction and magnitude of observed effects. To date, no study has compared how these factors jointly influence the use of semantic similarity measures to identify SSD and characterize dimensions of thought and language disturbance associated with SSD. Here, we sought to address this gap by systematically evaluating semantic similarity features derived from six language models across three levels of preprocessing and multiple comparison methods in a large sample of 228 participants. Our aims were: (1) to determine the optimal level of pre-processing for each language model in distinguishing SSD from healthy volunteers (HV), (2) to confirm the direction of diagnostic effects across semantic similarity metrics, and (3) to examine how these features relate to the dimensions of language disturbance described by impaired expressivity, inefficiency, and incoherence factors based on select items from The Scale for the Assessment of Thought, Language, and Communication (TLC) and SANS (3).

## 2. Methods

### 2.1. Participants and Recruitment

Participants were recruited from the outpatient and inpatient services at Zucker Hillside Hospital (Queens, NY) as part of three previously published studies, including Remora (n = 81), LPOP (n = 66), and ACES (n = 81). Previous studies have examined relationships between wide range of NLP metrics and clinical endpoints such as symptom severity, longitudinal illness trajectory and social cognition (5,8,34,35); the present study presents a new systematic evaluation of semantic similarities. The sample comprised 76 healthy volunteers and 152 individuals with schizophrenia spectrum disorders. All participants provided written informed consent. Study procedures were approved by the Institutional Review Board of the Feinstein Institutes for Medical Research.

### 2.2. Assessment and Diagnosis

Clinical diagnoses were established using standardized diagnostic interviews aligned with DSM-5 criteria. To evaluate dimensions of language disturbance, trained raters assessed 18 items from the Thought, Language, and Communication (TLC) scale (36) and 2 additional items from the Scale for the Assessment of Negative Symptoms (SANS) (37) that capture relevant aspects. Ratings were based on participants’ whole interviews, which included speech tasks as well as clinical assessments (BPRS, etc. write them here). Item ratings were used to compute three dimensions of language disturbance using previously published factor loadings with demonstrated cross-diagnostic validity: impaired expressivity, incoherence, and inefficiency, (3).

### 2.3. Speech Samples

Speech samples were elicited from each participant using open-ended prompts (e.g., “Tell me about yourself,” “How have you been recently?”) that were used for this study. Other tasks included picture description, phonemic and categorical fluency, etc., which were not utilized here. Recordings were transcribed verbatim with intuitive punctuations to define sentence boundaries. To protect participant privacy, prior to analysis, specific personal references were removed using manually annotated tags to replace personal identifiers with substitutes of the same function (e.g., “My sister Maria…” _→ “_My sister Jennifer…”). Non-speech verbalizations, such as symbolic placeholders for noises, laughter, unintelligible segments, and partial words, were also excluded from the transcripts. Final transcripts comprised turn-level dialogues in response to each individual prompt with speaker tags identifying the participant and the interviewer. Participants responded to one (N = 6) or two open-ended prompts (N = 222); when two transcripts (each in response to one of the above-mentioned open-ended prompts) were available for a participant, the average value of downstream semantic similarity features were used.

### 2.4. Pre-processing Levels

To compare varying degrees of textual pre-processing, we analyzed each transcript at three levels of pre-processing, each reflecting incremental removal of non-content words (Figure 1A).

1. Level 1 (Verbatim): Verbatim transcripts cleaned of non-verbal content as described above, but inclusive of disfluencies and filler words.
2. Level 2 (Disfluency-removed): Level 1 transcripts with additional removal of speech disfluencies, including filled pauses and repetitions.
3. Level 3 (Disfluency-removed and Stopword-filtered): Outputs from the embedding extraction applied to Level 2 were further filtered to remove stop words (i.e., non-content words, namely stop words, such as auxiliary verbs and prepositions). This sequencing allowed non-content words to provide contextual information to content-word embeddings prior to their elimination but removes them from the similarity calculation.

**Figure 1.**
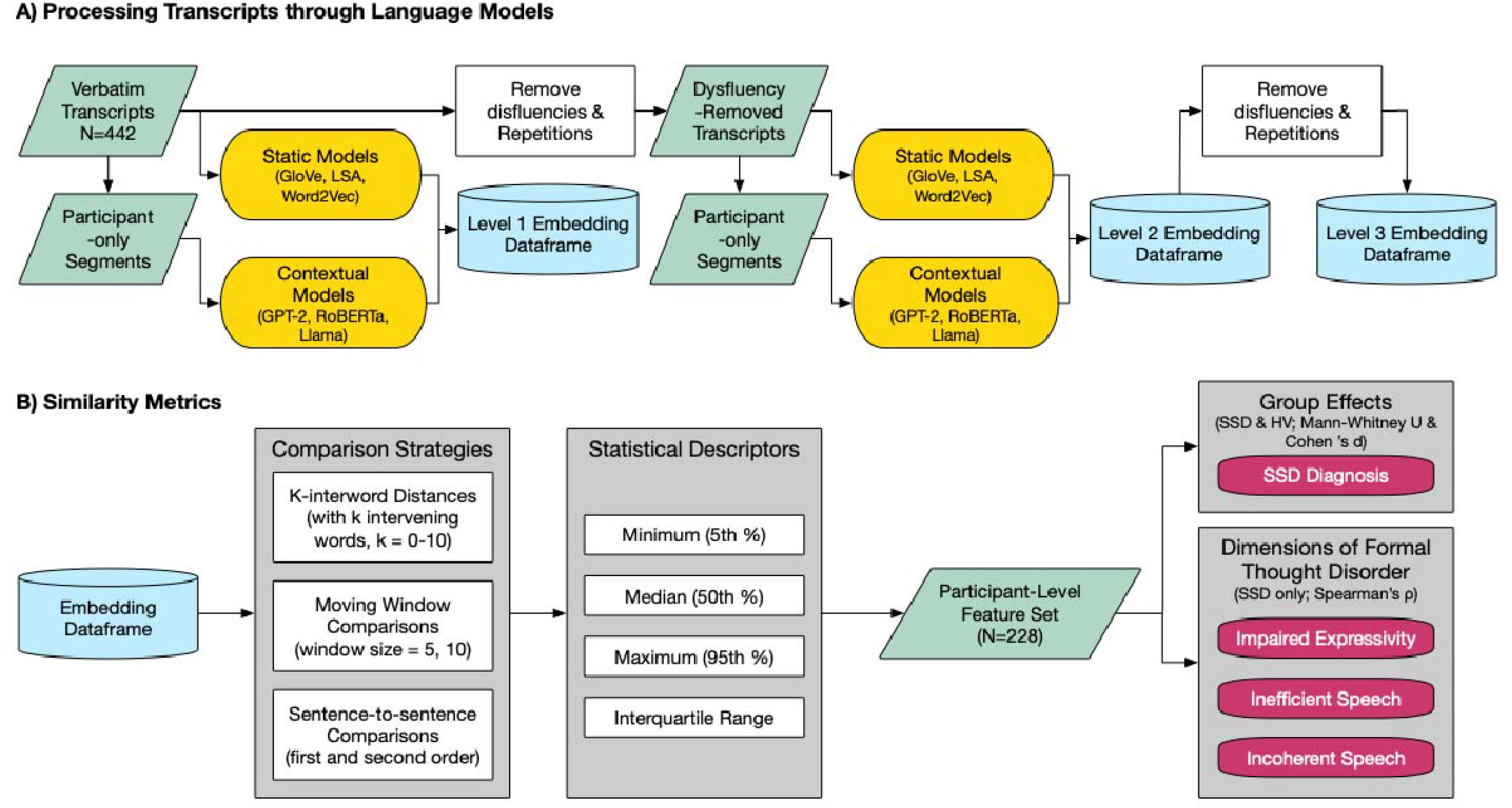
Pre-processing and Analytic Pipelines.

### 2.5. Embedding Extraction

Verbatim and disfluency-removed samples (Levels 1 and 2) underwent token-level embedding extraction using six pre-trained language models (Table 1): three non-contextual (GloVe, LSA, Word2Vec) and three contextual (GPT-2, RoBERTa, Llama). Models were selected based on their previous appearance in semantic similarity literature as well as their architectural variety. Of note, later versions of GPT are not open-weight models. Hence, GPT-2 was used in this study. For non-contextual models, interviewer speech was excluded and each word from the participant’s transcript was presented to the model independently. The corresponding embedding was recorded when existing in model’s corpus and skipped otherwise. For contextual models, full two-party conversations (participant and interviewer) were provided to the model to preserve discourse context. Token-level embeddings were extracted using each model’s native tokenizer. Tokens were mapped back to their originating words, and for words split into multiple tokens, the mean of the token embeddings was computed to yield a single word-level vector. After embedding extraction, interviewer turns were removed, retaining participant embeddings that benefited from the conversational context. Transcripts which exceeded the token limit of each model in length were parsed into overlapping segments and presented to the model separately. We avoided breaking sentences in the middle while keeping each segment with maximum allowed tokens. For tokens in the overlapping regions, the average embedding value was calculated.

**Table 1.** Language Model Characteristics. Non-contextual (static) models generate fixed embeddings for each word independent of surrounding text, whereas contextual models dynamically generate embeddings that vary as a function of linguistic context. Models included were Global Vectors for Word Representation (GloVe) (10), Latent Semantic Analysis (LSA) (11), Word2Vec (12), Robustly Optimized Bidirectional Encoder Representations from Transformers (RoBERTa) (15), Generative Pre-trained Transformer 2 (GPT-2) (13), and Large Language Model Meta AI (Llama) (49). Note: B = billion, M = million; K = thousand; GB = gigabyte.

| Model Name<br>(Version if applicable) | Contextualization | Architecture | Vector Dimension | Other Specifics |
| --- | --- | --- | --- | --- |
| **GloVe** | Non-contextual | Word co-occurrence matrix | 300 | Wikipedia 2014 and Gigaword 5 Corpus; 6B tokens; 400k vocabulary |
| **LSA** | Non-contextual | Word-document matrix | 300 | TASA educational corpus; ~38K text passages; 92K terms |
| **Word2Vec** | Non-contextual | Neural network | 300 | Google News corpus; ~100B words; 3M word and phrase vectors |
| **RoBERTa**<br>(RoBERTa Large) | Contextual | Bidirectional transformer encoder | 1024 | 355M Parameters; 24 layers; 160GB training corpus |
| **GPT-2**<br>(GPT2-XL) | Contextual | Unidirectional transformer decoder | 1600 | 1.5B parameters; 48 layers; 40GB corpus |
| **Llama**<br>(Hermes 3 Llama-3.1 8B) | Contextual | Unidirectional transformer decoder | 4096 | ~8B parameters; 32 layers |

### 2.6. Similarity Metrics

Word-level embeddings derived from participant speech at each of the three pre-processing level (1. Verbatim; 2. Disfluency-removed; and 3. Disfluency-removed and stopword-filtered), and each model (GloVe, LSA, Word2Vec, GPT-2, RoBERTa, and Llama) were used to compute the following similarity comparisons. Percentile and median-based statistics were chosen over mean, absolute minima and maxima to decrease sensitivity to outliers.

1. K-interword distances were generated by computing the cosine similarity between each word and the word occurring after k intervening words, for k = 0 to k=10. For example, the k-interword distance with k=0 compares a word to its immediate successor, whereas k=6 compares each word to the seventh successive word with six intervening words. For each transcript, the 5^th^ percentile (minimum), 95^th^ percentile (maximum), 50^th^ percentile (median), and interquartile range of all k-interword comparisons were calculated.
2. Moving-window similarity was computed with window widths of 5 and 10 words and a stride of one word. Within each window we computed the minimum, maximum, median, and interquartile range of the pairwise cosine similarities, and these statistics were subsequently averaged across all windows to produce sample-level measures.
3. Sentence embeddings were obtained by mean-pooling all word embeddings within each sentence at the given pre-processing level per standard practice in the field (38). Cosine similarity was then computed between consecutive sentences to assess first-order coherence and between sentences separated by one intervening sentence to assess second-order coherence. For each sample, the minimum, maximum, median, and interquartile range of these sentence-to-sentence similarity scores were calculated.

This procedure yielded 68 unique similarity metrics computed for each sample across three preprocessing levels and six embedding models, resulting in 1,224 similarity features per original sample (68 × 3 × 6; Figure 1B). For participants with two transcripts (N=222 participants), each feature was averaged across the two transcripts to obtain a single participant-level value. A complete list of semantic similarity features is provided in the appendix (Table S-1).

### 2.7. Statistical Analysis

Similarity features (n = 1,224) were compared between SSD and HV groups using the Mann–Whitney U test (39), with effect sizes summarized as Cohen’s d (d). The Mann-Whitney U test was chosen because it is robust to non-normally distributed data. To distinguish which pre-processing level optimizes group-level effect per model, we counted and reported number of significant features and maximum absolute effect size for each model–pre-processing level pair. Subsequently, for the dimensional analyses, we utilized the level of pre-processing at which each model yielded the highest number of significant features and largest effect sizes. For the dimensional analyses, we examined associations between features and latent dimensions of thought and language disturbance within the SSD group (3): impaired expressivity, incoherence, and inefficiency. Spearman’s rank correlation (_ρ)_ was reported.

Overall processing pipeline is schematically demonstrated in Figure 1. All statistical and NLP analyses, and data visualizations were conducted in Python 3 using pandas, SciPy, seaborn, and spaCy libraries (40–43).

## 3. Results

### 3.1. Participant Characteristics

Participants included 228 individuals (Table 2-A), comprising 152 participants with schizophrenia spectrum disorders (SSD) and 76 healthy volunteers (HV). The SSD and HV groups did not differ significantly in age (SSD: 27.9 ± 7.4 years; HV: 29.0 ± 6.8 years; *p* = 0.30) but differed in sex distribution (p = 0.02), with a higher proportion of males in the SSD group (64%) than in the HV group (46%). Race distribution did not differ significantly between groups (*p* = 0.14); across the full sample, 37% of participants identified as African American, 33% as Caucasian, 14% as Asian, and 16% as other race. As expected, clinical ratings of thought and language disturbance were significantly higher in the SSD group than in HVs, including TLC total score (SSD: 17 ± 15; HV: 2 ± 3; *p* < 0.001) and all three latent language-disturbance factors: inefficiency, incoherence, and impaired expressivity (all *p* < 0.001)

**Table 2.** Participant Characteristics. Overall and group-wise participant characteristics are summarized. Group differences were assessed using t-tests for continuous variables (age, TLC scores, and speech sample characteristics) and chi-square tests for categorical variables (sex and race).

|  | Overall | SSD | HV | p Value |
| --- | --- | --- | --- | --- |
| **A. Participant Characteristics** |  |  |  |  |
| **N** | 228 | 152 | 76 |  |
| **Age** |  | 27.9 ± 7.4 | 29.0 ± 6.8 | 0.30 |
| **Sex** |  |  |  | 0.02 |
| **Female** | 96 (42%) | 55 (36%) | 41 (54%) |  |
| **Male** | 132 (58%) | 97 (64%) | 35 (46%) |  |
| **Race** |  |  |  | 0.14 |
| **African American** | 85 (37%) | 58 (38%) | 27 (35.5%) |  |
| **Asian** | 31 (14%) | 19 (12.5%) | 12 (16%) |  |
| **Caucasian** | 75 (33%) | 45 (29.5%) | 30 (39.5%) |  |
| **Other** | 37 (16%) | 30 (20%) | 7 (9%) |  |
| **TLC Total Score** |  | 17 ± 15 | 2 ± 3 | < 0.001 |
| **Inefficiency Factor** |  |  |  | < 0.001 |
| **Incoherence Factor** |  |  |  | < 0.001 |
| **Impaired Expressivity Factor** |  |  |  | < 0.001 |
| **B. Sample Characteristics** |  |  |  |  |
| **Number of words** | 145 (159) | 143 (168) | 149 (138) | 0.69 |
| **Number of sentences** | 12 (11) | 12 (12) | 12 (10) | 0.91 |
| **Number of stopwords** |  | 80 (95) | 79 (77) | 0.88 |
| **Repetitions %** |  | 1.38 (2.64) | 1.16 (1.99) | 0.33 |
| **Filled pauses %** |  | 5.43 (5.77) | 6.53 (4.97) | 0.04 |

### 3.2. Speech Sample Description

Altogether, 450 documents were available from 228 participants, each corresponding to one of two open-ended prompts described above (see Table 2-B). Eight samples containing fewer than 10 words were excluded from analysis (7 from individuals with SSD and 1 from an HV participant). On average, samples comprised 145 words across 12 utterances. The SSD and HV groups did not differ significantly in the number of words per sample (SSD: 143; HV: 149; *p* = 0.69), number of utterances (both 12; *p* = 0.91), number of stopwords (SSD: 80; HV: 79; *p* = 0.88), or repetition rate (SSD: 1.38% of total words; HV: 1.16% of total words; *p* = 0.33). Healthy volunteers used filled pauses more frequently than participants with SSD (HV: 6.53% of total words vs. SSD: 5.43% of total words) with statistical significance (*p* = 0.04).

### 3.3. Effect of Pre-processing on Semantic Similarity Group Effects

The effect of pre-processing on the group effects of semantic similarity features in terms of number of features reaching p < 0.05 the maximum observed effect size (d) is illustrated in Figure 2. As non-content words were progressively removed, models showed distinct trends in terms of their maximum effect size and total number of significant features. GPT-2 exhibited an incremental performance increase; RoBERTa, GloVe, and Word2Vec showed decreased performance; and LSA and Llama were comparatively insensitive to pre-processing. For GPT-2, further pre-processing particularly enhanced signal between SSD diagnosis and increased minimum and median semantic similarity in k-interword, moving-window, and sentence-to-sentence similarity comparisons. For Word2Vec same minimum and median semantic similarity signal diminished with pre-processing but only in word and window comparisons. For RoBERTa and GloVe, no generalizable pattern was observed (Table-S2).

**Figure 2.**
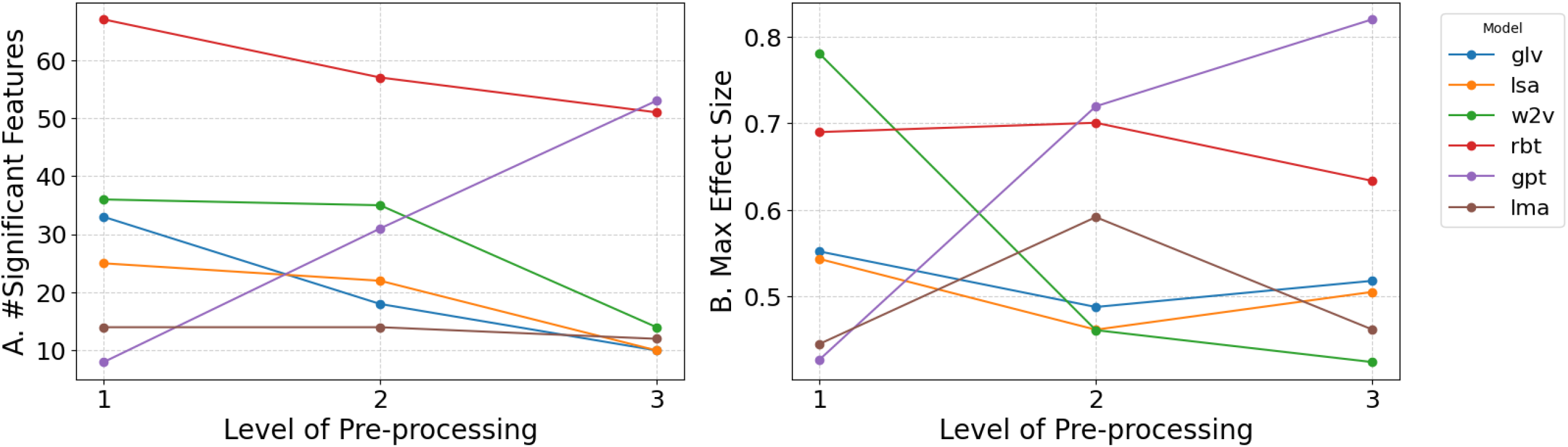
Model performance across preprocessing levels. Model performance was evaluated at preprocessing Level 1 (verbatim), Level 2 (disfluency-removed), and Level 3 (disfluency-removed and stopword-filtered) according to (A) the number of semantic similarity features significantly associated with SSD diagnosis (p < 0.05) and (B) the maximum observed effect size (Cohen’s d) among those features. Note: glv = GloVe; lsa = LSA; w2v = Word2Vec; rbt = RoBERTa; gpt = GPT-2; lma = Llama.

RoBERTa at pre-processing level 1 outperformed all other model–pre-processing combinations in the number of significant features, with 67 features. GPT-2 at pre-processing level 3 achieved the highest effect size of 0.82 among all comparisons. The best pre-processing level for each model was selected for further analysis. They included GloVe at pre-processing level 1, LSA at pre-processing level 1, Word2Vec at pre-processing level 1, RoBERTa at pre-processing level 1, GPT-2 at pre-processing level 3, and Llama at pre-processing level 2. Comprehensive statistics for all comparisons (68 features × six models × three pre-processing levels) are provided in the supplement (Table S-2).

### 3.4. Association with Diagnosis across Language Models

Feature-wide association of SSD diagnosis with semantic similarity metrics derived from the six models at their optimized level of pre-processing are illustrated in Figure 3. Selected features including even k-words are included here for better visualization (See Figure 1-S for all comparisons). Overall, SSD diagnosis was associated with increased semantic similarity across models, pre-processing approaches and statistical metrics, with the exception of inter-quartile range comparisons.

**Figure 3.**
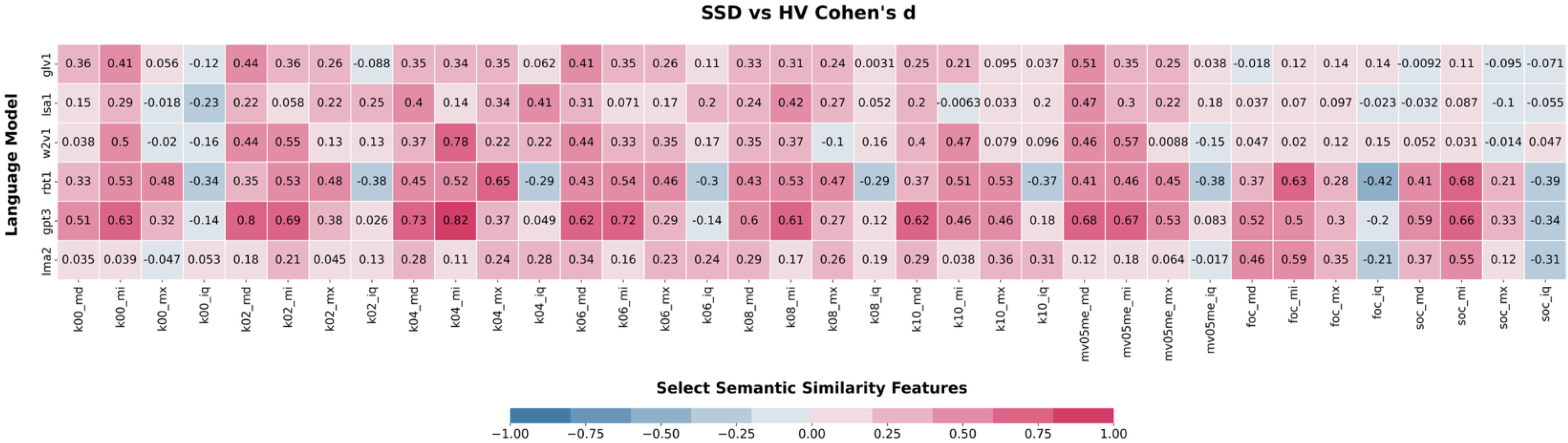
Associations with SSD diagnosis at optimal level of preprocessing. Associations between SSD diagnosis and semantic similarity metrics (mi: minimum, md: median, mx: maximum, iq: interquartile range) are shown for language models at their respective optimized pre-processing level. Similarity measures include k-interword comparisons, moving-window metrics, and first- and second-order sentence-to-sentence similarity (See Table S-1 in supplements for feature-wide descriptions). Effects are summarized as Cohen’s d from Mann–Whitney U. Note: glv1 = GloVe at pre-processing level 1; lsa1 = LSA at pre-processing level 1, w2v1 = Word2Vec at pre-processing level 1, rbt1 = RoBERTa at pre-processing level 1, gpt3 = GPT-2 at pre-processing level 1, lma2 = Llama at pre-processing level 2.

Within k-interword analyses, GPT-2 outperformed other combinations, showing consistently strong associations between SSD and increased *minimum* and *median* interword similarities across all lags k0-k10 (d = 0.46–0.82). RoBERTa produced comparatively stronger signals with increased *maximum* interword similarity being associated with SSD (d = 0.46–0.69). Llama yielded weaker effects overall, with gradual improvement at more distant lags and its highest effect size observed for increased maximum similarity at k-10 (d = 0.36). Non-contextual models also showed multiple significant associations with SSD diagnosis, but without a consistent pattern across interword distance or statistical summary; notably, Word2Vec achieved the highest effect size among non-contextual models for increased *minimum* interword similarity at the 4-word interval (d = 0.78).

Among moving window comparisons, effect sizes were generally stable within each model across window size (five vs. 10 words) and statistical approach. GPT-2 was the best performing combination, with multiple features at both moving-window sizes reaching d _≈ 0_.8. Llama was the weakest, showing minimal improvement with larger windows (from d _≈ 0_.12–0.18 to d _≈ 0_.21–0.22). RoBERTa, LSA, GloVe, and Word2Vec produced features with peak effect sizes in d _≈ 0_.4–0.6 range.

For first- and second-order sentence-to-sentence similarity, none of the non-contextual models (LSA, Glove, Word2Vec) yielded significant associations with SSD diagnosis. In contrast, all three contextual models (GPT-2, RoBERTa, and Llama) did demonstrate significant group effects, where SSD was associated with increased *minimum*, *median*, and *maximum* sentence-to-sentence similarity and decreased interquartile range.

### 3.5. Association with Dimensions of Thought and Language Disturbance

Figure 4 presents feature-wide correlations between similarity metrics from each model and three language disturbance factors, namely, inefficiency (Figure 4-A), and incoherence (Figure 4-B), and impaired expressivity (Figure 4-C). Overall, all models contributed features with the strongest performance in _ρ =_ 0.2–0.35 range, with distinct patterns in the number of significant correlations across combination as described below.

**Figure 4.**
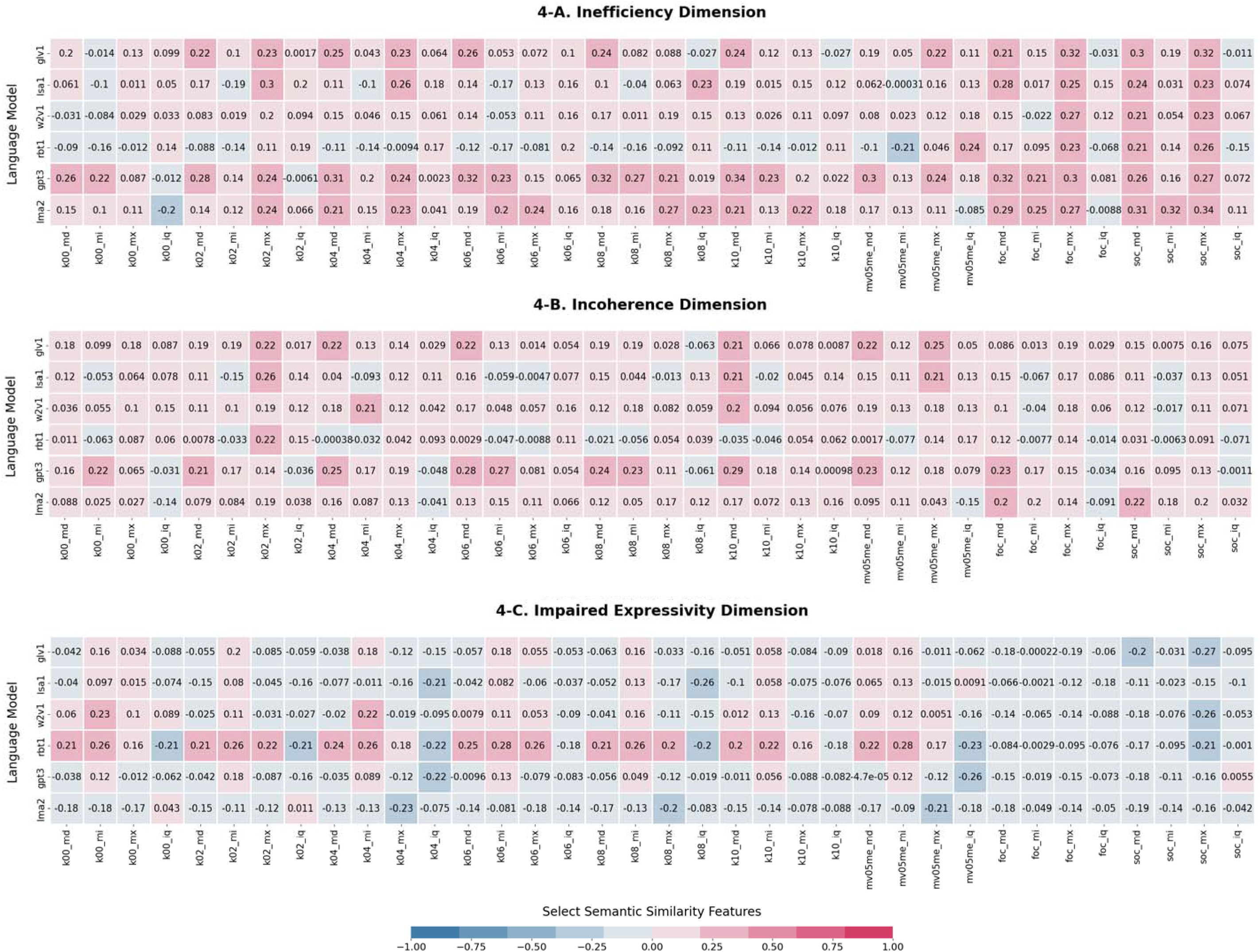
Semantic similarity feature associations with language-disturbance dimensions across language models at their optimized level of preprocessing. Select feature-wide associations between semantic similarity metrics (mi: minimum, md: median, mx: maximum, iq: interquartile range) and language-disturbance dimensions are shown for language models at their respective optimized preprocessing level. Similarity measures include k-interword comparisons, moving-window metrics, and first- and second-order sentence-to-sentence similarity (see Table S-1 in the Supplement for feature descriptions). Associations are summarized as Spearman’s _ρ._ Panels correspond to inefficiency (Panel 4-A), incoherence (Panel 4-B), and impaired expressivity (Panel 4-C) dimensions. Note: glv1 = GloVe at pre-processing level 1; lsa1 = LSA at pre-processing level 1, w2v1 = Word2Vec at pre-processing level 1, rbt1 = RoBERTa at pre-processing level 1, gpt3 = GPT-2 at pre-processing level 3, lma2 = Llama at pre-processing level 2.

GPT-2 produced the greatest number of significant correlations with the inefficiency factor. Greater inefficiency was associated with increased similarity across k-interword, moving-window, and sentence-to-sentence metrics. Increase in *maximum* first-order sentence-to-sentence similarity, as well as *median* and *maximum* second-order similarity, were correlated with greater inefficiency across all six models.

GPT-2 also outperformed other models in correlation with incoherence. More incoherent speech was associated with increased *minimum* and *median* similarity in multiple k-interword and moving-window comparisons and decreased average interquartile range in 5- and 10-word windows.

RoBERTa yielded the greatest number of significant correlations with impaired expressivity, characterized by increased *minimum*, *median*, and *maximum* similarity and decreased *interquartile range* with k-interword and moving-window metrics tracking with severity. Higher median and maximum sentence-to-sentence similarities were correlated with lower impaired expressivity in RoBERTa, GloVe and Word2Vec. Few sporadic significant associations observed between impaired expressivity and semantic similarity features extracted from other language models.

## 4. Discussion

In this study, we systematically examined how semantic similarity features derived from multiple embedding models and preprocessing levels relate to SSD diagnosis and to dimensions of thought and language disturbance. Three main findings emerged: 1. SSD diagnosis and dimensions of thought and language disturbance were generally associated with *increased* semantic similarity across models and comparisons; 2. Different dimensions of thought and language disturbance had distinct semantic similarity fingerprints; and 3. Language models exhibited varying behavior in response to preprocessing of the transcribed text.

A central finding of this study was that the existence of SSD was associated with higher semantic similarity and lower dispersion across models and comparison approaches: average, minimum and maximum semantic similarity is SSD was broadly increased with respect to healthy controls across word, window, and sentence-level analyses, and this increase was commonly accompanied by reduction in semantic variability as reflected in decreased inter-quartile range. This pattern aligns closely with recent work interpreting elevated similarity as evidence of a constricted semantic space (29,32). In addition, as increased semantic similarity found to be associated with both positive and negative dimensions of thought disorder, our results indicate that this computational metric is relevant not only to impoverishment in SSD but also to disorganization. Prior studies indicated that increased semantic similarity in SSD is a product of word choice that occurs from tightly clustered semantic spaces, rather than being merely a result of insufficient diversity, i.e., using fewer instances of words to build discourse (26). Taken together, our findings indicate that disorganization is not always the result of co-occurrence of semantically unrelated words or sentences in speech; rather, it may also result from juxtaposition of semantically similar words or sentences in incoherent or inefficient manners. In addition to future informing efforts to develop objective speech and language markers for psychosis, these findings may also guide future work in synthetic linguistic data representing formal thought disorder using generative language models (44).

In the present data, increased sentence-level similarity was more prominently related to inefficient speech (e.g., tangentiality and circumstantiality), compared to incoherence. This finding is in line with the previous conceptualization of inefficiency as disruption of global meaning intent that is carried between sentences, in contrast to incoherence, which reflects disruption of local meaning that occurs within sentences (3). This association with *increased* similarity may reflect the fact that rather than semantic divergence (18), inefficient speech often remains semantically close across sentences because it elaborates redundantly, circles around a topic, or fails to advance discourse efficiently. In contrast, incoherence may be better captured by shorter-range metrics, such as local interword or moving-window comparisons, where disruptions or over-constraint in immediate semantic transitions are more visible. More broadly, these findings support the idea that different dimensions of thought and language disturbance in SSD may operate at different linguistic scales and units, and that embedding-based measures should be interpreted in relation to the level of discourse they quantify.

Preprocessing had substantial differential effects on the relationship between downstream semantic similarity measurements and clinical measures. RoBERTa showed the strongest overall performance on minimally processed transcripts, whereas GPT-2 performed best after the most extensive preprocessing. This suggests that, in embedding methodology, depending on language model selection, pre-processing might preserve or attenuate linguistic signals relevant to clinical outcomes. For RoBERTa, minimally processed speech may retain discourse scaffolding carried by function words, repetitions, and disfluencies that remain clinically informative, particularly for impoverishment-related or expressive deficits. By contrast, GPT-2 may benefit from removal of non-content words because this sharpens sequential relations among content-bearing words, thereby enhancing signals relevant to disorganization. This different pattern could also be explained in terms of models’ intrinsic architecture: In general, RoBERTa’s bidirectional text processing might be robust to repetitions, disfluencies, etc., whereas, GPT-2 is trained and optimized to process text unidirectionally to predict the next word, as such, unstructured, conversational “noise” might disrupt its prediction flow and requires aggressive cleaning. Finally, this finding is also consistent with our previous hypothesis about semantic localization of language disturbance in psychosis, namely that disorganization-related signals are more concentrated in the semantic core of speech (i.e., information related to events and their participants), whereas impaired expressivity is more diffusely distributed across core and peripheral structure (i.e., information related to setting, manner, etc.) (5,45).

Several limitations should be noted. Although we compared six embedding models, we did not include representations from some newer or more advanced models and model variants. For example, at the time of this study, token embeddings were only readily extractable from GPT-2 among OpenAI models; also, more powerful models were not necessarily open source or open weight for us to safely extract model internal representations. However, our results indicated that despite the overall superiority of more advanced models, older non-contextual language models were still able to generate features with effect sizes comparable to those of large language models despite not directly encoding the syntactic information in the text or discourse context from the interviewer. Moreover, the models tested in our work are relatively light and cost-efficient to deploy and reproduce, requiring much less compute compared to the more advanced large language models. In addition, our analyses were based on open-ended interview speech, and the findings may not generalize directly to more constrained elicitation tasks such as picture description or verbal fluency. To ensure methodological consistency across contextual and non-contextual models, we constructed sentence-level embeddings by mean-pooling the embeddings of their constituent words. However, this approach does not fully leverage the capabilities of advanced contextual models, which can generate sentence-level representations informed by the broader conversational context. Future studies may explore such context-aware embeddings to better capture semantic relationships across sentences. Finally, the observed relationships with language-disturbance factors were modest in magnitude, indicating that semantic similarity captures only certain aspects of clinically rated language abnormality. Other aspects may be better characterized by complementary metrics such as perplexity (46) or alternative computational approaches such as semantic graph representation of discourse structure (47) or measures of referential coherence (48). Integrating these measures may provide a more comprehensive characterization of language disturbance in SSD.

## 5. Conclusion

Semantic similarity alteration in SSD was characterized predominantly by higher similarity and reduced variability across models, feature types, and pre-processing approaches. RoBERTa performed best on minimally processed transcripts and was most informative for impaired expressivity, whereas GPT-2 performed best after maximal preprocessing and was most informative for inefficiency and incoherence, shedding practical insights on the utility and selection of language models in future SSD research. These findings suggest that negative formal thought disorder may be better captured with minimal preprocessing with bidirectional approach, which is sensitive to broader speech context, whereas disorganization-related signals may emerge more clearly after stronger preprocessing with unidirectional approach agnostic to downstream context. Furthermore, inefficiency was more prominently associated with sentence-level similarity, whereas incoherence was more closely related to shorter-range interword and moving-window comparisons, suggesting that these dimensions of language disturbance may preferentially reflect disruptions in global versus local semantic organization, respectively.

## Data Availability

All data produced in the present study are available upon reasonable request to the authors.

## Acknowledgements

We thank the participants for their contributions and the following research coordinators for their help in data collection: Sarah A. Berretta, Leily M. Behbehani. Funding was received from K23 MH130750 (SXT) and the Brain and Behavior Research Foundation Young Investigator Grant (SXT). Departmental funding from the Feinstein Institutes for Medical Research was used to fund data collection on the Remora sample. Winterlight Labs provided funding for data collection on the LPOP and ACES samples, but did not play a role in the analytical planning, analysis, interpretation or drafting of this manuscript.

## References

1. Kuperberg GR (2008): Building meaning in schizophrenia. Clin EEG Neurosci 39: 99–102.

2. Kircher T, Bröhl H, Meier F, Engelen J (2018): Formal thought disorders: from phenomenology to neurobiology. Lancet Psychiatry 5: 515–526.

3. Tang SX, Hänsel K, Cong Y, Nikzad AH, Mehta A, Cho S, et al. (2023): Latent Factors of Language Disturbance and Relationships to Quantitative Speech Features. Schizophr Bull 49: S93–S103.

4. Andreasen NC (1982): Negative symptoms in schizophrenia. Definition and reliability. Arch Gen Psychiatry 39: 784–788.

5. Nikzad AH, Cong Y, Berretta S, Hänsel K, Cho S, Pradhan S, et al. (2022): Who does what to whom? graph representations of action-predication in speech relate to psychopathological dimensions of psychosis. Schizophrenia 8: 58.

6. Palominos C, Kirdun M, Nikzad AH, Spilka MJ, Homan P, Sommer IE, et al. (2025): A single composite index of semantic behavior tracks symptoms of psychosis over time. Schizophr Res 279: 116–127.

7. Tang SX, Kriz R, Cho S, Park SJ, Harowitz J, Gur RE, et al. (2021): Natural language processing methods are sensitive to sub-clinical linguistic differences in schizophrenia spectrum disorders. Npj Schizophr 7: 25.

8. Tang SX, Cong Y, Nikzad AH, Mehta A, Cho S, Hänsel K, et al. (2023): Clinical and computational speech measures are associated with social cognition in schizophrenia spectrum disorders. Schizophr Res 259: 28–37.

9. Parola A, Lin JM, Simonsen A, Bliksted V, Zhou Y, Wang H, et al. (2023): Speech disturbances in schizophrenia: Assessing cross-linguistic generalizability of NLP automated measures of coherence. Schizophr Res 259: 59–70.

10. Pennington J, Socher R, Manning C (2014): GloVe: Global Vectors for Word Representation. In: Moschitti A, Pang B, Daelemans W, editors. Proceedings of the 2014 Conference on Empirical Methods in Natural Language Processing (EMNLP). Doha, Qatar: Association for Computational Linguistics, pp 1532–1543.

11. Foltz PW (1996): Latent semantic analysis for text-based research. Behav Res Methods Instrum Comput 28: 197–202.

12. Mikolov T, Chen K, Corrado G, Dean J (2013, September 7): Efficient Estimation of Word Representations in Vector Space [no. arXiv:1301.3781]. arXiv. 10.48550/arXiv.1301.3781

13. Radford A, Wu J, Child R, Luan D, Amodei D, Sutskever I (2019): Language Models are Unsupervised Multitask Learners.

14. Touvron H, Lavril T, Izacard G, Martinet X, Lachaux M-A, Lacroix T, et al. (2023, February 27): LLaMA: Open and Efficient Foundation Language Models [no. arXiv:2302.13971]. arXiv. 10.48550/arXiv.2302.13971

15. Liu Y, Ott M, Goyal N, Du J, Joshi M, Chen D, et al. (2019, July 26): RoBERTa: A Robustly Optimized BERT Pretraining Approach [no. arXiv:1907.11692]. arXiv. 10.48550/arXiv.1907.11692

16. Corcoran CM, Cecchi GA (2020): Using Language Processing and Speech Analysis for the Identification of Psychosis and Other Disorders. Biol Psychiatry Cogn Neurosci Neuroimaging 5: 770–779.

17. Elvevåg B, Foltz PW, Weinberger DR, Goldberg TE (2007): Quantifying incoherence in speech: an automated methodology and novel application to schizophrenia. Schizophr Res 93: 304–316.

18. Morgan SE, Diederen K, Vértes PE, Ip SHY, Wang B, Thompson B, et al. (2021): Natural Language Processing markers in first episode psychosis and people at clinical high-risk. Transl Psychiatry 11: 630.

19. Lundin NB, Jones MN, Myers EJ, Breier A, Minor KS (2022): Semantic and phonetic similarity of verbal fluency responses in early-stage psychosis. Psychiatry Res 309: 114404.

20. Pauselli L, Halpern B, Cleary SD, Ku BS, Covington MA, Compton MT (2018): Computational linguistic analysis applied to a semantic fluency task to measure derailment and tangentiality in schizophrenia. Psychiatry Res 263: 74–79.

21. Cho S, Cong Y, Mehta A, Nikzad AH, Berretta SA, Behbehani LM, et al. (2026): Unique signatures in verbal fluency task performance in schizophrenia and depression. Schizophr Res Cogn 43: 100407.

22. Corcoran CM, Carrillo F, Fernández-Slezak D, Bedi G, Klim C, Javitt DC, et al. (2018): Prediction of psychosis across protocols and risk cohorts using automated language analysis. World Psychiatry 17: 67–75.

23. Bedi G, Carrillo F, Cecchi GA, Slezak DF, Sigman M, Mota NB, et al. (2015): Automated analysis of free speech predicts psychosis onset in high-risk youths. Npj Schizophr 1: 15030.

24. Dalal TC, Park MTM, Silva AM, Iskhakova S, Voppel A, Brierley NJ, et al. (2026): Clinical psychopathology-based early relapse prediction model using speech and language in psychosis. Schizophr Res Cogn 43: 100392.

25. Alonso-Sánchez MF, Ford SD, MacKinley M, Silva A, Limongi R, Palaniyappan L (2022): Progressive changes in descriptive discourse in First Episode Schizophrenia: a longitudinal computational semantics study. Schizophrenia 8: 36.

26. Alonso-Sánchez MF, Limongi R, Gati J, Palaniyappan L (2023): Language network self-inhibition and semantic similarity in first-episode schizophrenia: A computational-linguistic and effective connectivity approach. Schizophr Res 259: 97–103.

27. Çabuk T, Sevim N, Mutlu E, Yağcıoğlu AEA, Koç A, Toulopoulou T (2024): Natural language processing for defining linguistic features in schizophrenia: A sample from Turkish speakers. Schizophr Res 266: 183–189.

28. Çokal D, Aloraini A, Palominos CF, Demirlek C, Verim B, Yalınçetin B, et al. (2025): Three dimensions of speech coherence in people with early psychosis and their family members. Schizophrenia 12: 2.

29. He R, Palominos C, Zhang H, Alonso-Sánchez MF, Palaniyappan L, Hinzen W (2024): Navigating the semantic space: Unraveling the structure of meaning in psychosis using different computational language models. Psychiatry Res 333: 115752.

30. Palominos C, He R, Fröhlich K, Mülfarth RR, Seuffert S, Sommer IE, et al. (2024): Approximating the semantic space: word embedding techniques in psychiatric speech analysis. Schizophrenia 10: 114.

31. He R, Grodzki R, Altay N, Aston B, Alonso-Sánchez MF, Homan P, et al. (2026): Reduced linguistic coherence in psychosis defies semantic similarity accounts and relates to altered large-scale cortical hierarchy. Sci Rep 16: 7799.

32. Palominos C, Stein F, Kircher T, Ayesa-Arriola R, Palaniyappan L, Homan P, et al. (2025): Lexical meaning is lower dimensional in psychosis. Sci Rep 16: 859.

33. Ciampelli S, de Boer JN, Koops S, Troelstra E, Jebens A, Marsman J-BC, et al. (2026): Automated Speech-Based Modeling of Item-Level Symptom Severity in Schizophrenia. JAMA Netw Open 9: e2620239.

34. Mehta A, Nikzad AH, Cong Y, Cho S, Pradhan S, Tang SX (2025): Sentiment in speech is associated with symptom severity in psychosis. Cognit Neuropsychiatry 30: 199–210.

35. Tang SX, Li J, Brosch K, Massouh N, Yin S, Bhola S, et al. (2026): ACES: ascertaining diagnosis classification with elicited speech in individuals with heterogeneous, comorbid psychiatric disorders. Psychiatry Res 365: 117389.

36. Andreasen NC (1986): Scale for the assessment of thought, language, and communication (TLC). Schizophr Bull 12: 473–482.

37. Andreasen NC (1989): The Scale for the Assessment of Negative Symptoms (SANS): conceptual and theoretical foundations. Br J Psychiatry Suppl 49–58.

38. Tunstall L, von Werra L, Wolf T (2022): Natural Language Processing with Transformers, Revised Edition. O’Reilly Media. Retrieved September 28, 2026, from https://www.oreilly.com/library/view/natural-language-processing/9781098136789/

39. Mann HB, Whitney DR (1947): On a Test of Whether one of Two Random Variables is Stochastically Larger than the Other. Ann Math Stat 18: 50–60.

40. McKinney W (2011): pandas: a Foundational Python Library for Data Analysis and Statistics. Retrieved July 26, 2026, from https://www.semanticscholar.org/paper/pandas%3A-a-Foundational-Python-Library-for-Data-and-McKinney/1a62eb61b2663f8135347171e30cb9dc0a8931b5

41. Virtanen P, Gommers R, Oliphant TE, Haberland M, Reddy T, Cournapeau D, et al. (2020): SciPy 1.0: fundamental algorithms for scientific computing in Python. Nat Methods 17: 261–272.

42. Fantechi A, Gnesi S, Livi S, Semini L (2021): A spaCy-based tool for extracting variability from NL requirements. Proceedings of the 25th ACM International Systems and Software Product Line Conference - Volume B 32–35.

43. Waskom ML (2021): seaborn: statistical data visualization. J Open Source Softw 6: 3021.

44. Fradkin I, Nour MM, Dolan RJ (2023): Theory-Driven Analysis of Natural Language Processing Measures of Thought Disorder Using Generative Language Modeling. Biol Psychiatry Cogn Neurosci Neuroimaging 8: 1013–1023.

45. Nikzad AH, Cong Y, Berretta S, Behbehani L, Pradhan S, Tang S (2023): Disorganization Disrupts the Core: Localizing Clinical Dimensions of Speech and Language Disturbance Within Clause Structure Using Multi-Layered Semantic Graph Analysis. NEUROPSYCHOPHARMACOLOGY, vol. 48 48: 362–362.

46. Xu W, Pakhomov S, Heagerty P, Horvitz E, Bradley ER, Woolley J, et al. (2025): Perplexity and proximity: Large language model perplexity complements semantic distance metrics for the detection of incoherent speech. J Biomed Inform 170: 104899.

47. Nikzad AH, Lysaker PH, Minor KS, Leonhardt B, Liberman MY, Vohs J, et al. (2025): Evolution of Linguistic Markers of Agency, Centrality and Content During Metacognitive Therapy for Psychosis: A Pilot Exploratory Study. Early Interv Psychiatry 19: e13628.

48. Palominos C, Figueroa-Barra A, Hinzen W (2023): Coreference Delays in Psychotic Discourse: Widening the Temporal Window. Schizophr Bull 49: S153–S162.

49. Teknium R, Quesnelle J, Guang C (2024, August 15): Hermes 3 Technical Report [no. arXiv:2408.11857]. arXiv. 10.48550/arXiv.2408.11857

