## Supplementary_Material for "Systematic evaluation of semantic similarity across language models for schizophrenia spectrum disorders"

**Table S-1.** Feature-wide nomenclature, characterization, and definition of semantic similarity metrics.

| Feature name | Similarity Comparison | Summary Statistics | Definition |
| --- | --- | --- | --- |
| k00_md | 0-interword | Median | Median of K-0 cosine similarity distances |
| k00_mi | 0-interword | Minimum | Minimum of K-0 cosine similarity distances |
| k00_mx | 0-interword | Maximum | Maximum of K-0 cosine similarity distances |
| k00_iq | 0-interword | Interquartile range | Interquartile range of K-0 cosine similarity distances |
| k01_md | 1-interword | Median | Median of K-1 cosine similarity distances |
| k01_mi | 1-interword | Minimum | Minimum of K-1 cosine similarity distances |
| k01_mx | 1-interword | Maximum | Maximum of K-1 cosine similarity distances |
| k01_iq | 1-interword | Interquartile range | Interquartile range of K-1 cosine similarity distances |
| k02_md | 2-interword | Median | Median of K-2 cosine similarity distances |
| k02_mi | 2-interword | Minimum | Minimum of K-2 cosine similarity distances |
| k02_mx | 2-interword | Maximum | Maximum of K-2 cosine similarity distances |
| k02_iq | 2-interword | Interquartile range | Interquartile range of K-2 cosine similarity distances |
| k03_md | 3-interword | Median | Median of K-3 cosine similarity distances |
| k03_mi | 3-interword | Minimum | Minimum of K-3 cosine similarity distances |
| k03_mx | 3-interword | Maximum | Maximum of K-3 cosine similarity distances |
| k03_iq | 3-interword | Interquartile range | Interquartile range of K-3 cosine similarity distances |
| k04_md | 4-interword | Median | Median of K-4 cosine similarity distances |
| k04_mi | 4-interword | Minimum | Minimum of K-4 cosine similarity distances |
| k04_mx | 4-interword | Maximum | Maximum of K-4 cosine similarity distances |
| k04_iq | 4-interword | Interquartile range | Interquartile range of K-4 cosine similarity distances |
| k05_md | 5-interword | Median | Median of K-5 cosine similarity distances |
| k05_mi | 5-interword | Minimum | Minimum of K-5 cosine similarity distances |
| k05_mx | 5-interword | Maximum | Maximum of K-5 cosine similarity distances |
| k05_iq | 5-interword | Interquartile range | Interquartile range of K-5 cosine similarity distances |
| k06_md | 6-interword | Median | Median of K-6 cosine similarity distances |
| k06_mi | 6-interword | Minimum | Minimum of K-6 cosine similarity distances |
| k06_mx | 6-interword | Maximum | Maximum of K-6 cosine similarity distances |
| k06_iq | 6-interword | Interquartile range | Interquartile range of K-6 cosine similarity distances |
| k07_md | 7-interword | Median | Median of K-7 cosine similarity distances |
| k07_mi | 7-interword | Minimum | Minimum of K-7 cosine similarity distances |
| k07_mx | 7-interword | Maximum | Maximum of K-7 cosine similarity distances |
| k07_iq | 7-interword | Interquartile range | Interquartile range of K-7 cosine similarity distances |
| k08_md | 8-interword | Median | Median of K-8 cosine similarity distances |
| k08_mi | 8-interword | Minimum | Minimum of K-8 cosine similarity distances |
| k08_mx | 8-interword | Maximum | Maximum of K-8 cosine similarity distances |
| k08_iq | 8-interword | Interquartile range | Interquartile range of K-8 cosine similarity distances |
| k09_md | 9-interword | Median | Median of K-9 cosine similarity distances |
| k09_mi | 9-interword | Minimum | Minimum of K-9 cosine similarity distances |
| k09_mx | 9-interword | Maximum | Maximum of K-9 cosine similarity distances |
| k09_iq | 9-interword | Interquartile range | Interquartile range of K-9 cosine similarity distances |
| k10_md | 10-interword | Median | Median of K-10 cosine similarity distances |
| k10_mi | 10-interword | Minimum | Minimum of K-10 cosine similarity distances |
| k10_mx | 10-interword | Maximum | Maximum of K-10 cosine similarity distances |
| k10_iq | 10-interword | Interquartile range | Interquartile range of K-10 cosine similarity distances |
| mv05me_md | Median in MV-5 | Mean | Mean of median cosine similarity distances with 5-word window size and 1-word stride |
| mv05me_mi | Minimum in MV-5 | Mean | Mean of minimum cosine similarity distances with 5-word window size and 1-word stride |
| mv05me_mx | Maximum in MV-5 | Mean | Mean of maximum cosine similarity distances with 5-word window size and 1-word stride |
| mv05me_iq | Interquartile range in MV-5 | Mean | Mean of interquartile range of cosine similarity distances with 5-word window size and 1-word stride |
| mv10me_md | Median in MV-10 | Mean | Mean of median cosine similarity distances with 10-word window size and 1-word stride |
| mv10me_mi | Minimum in MV-10 | Mean | Mean of minimum cosine similarity distances with 10-word window size and 1-word stride |
| mv10me_mx | Maximum in MV-10 | Mean | Mean of maximum cosine similarity distances with 10-word window size and 1-word stride |
| mv10me_iq | Interquartile range in MV-10 | Mean | Mean of interquartile range of cosine similarity distances with 10-word window size and 1-word stride |
| foc_md | First-order coherence | Median | Median cosine similarity between consecutive sentences |
| foc_mi | First-order coherence | Minimum | Minimum cosine similarity between consecutive sentences |
| foc_mx | First-order coherence | Maximum | Maximum cosine similarity between consecutive sentences |
| foc_iq | First-order coherence | Interquartile range | Interquartile range of cosine similarity between consecutive sentences |
| soc_md | Second-order coherence | Median | Median cosine similarity between sentences separated by one intervening sentence |
| soc_mi | Second-order coherence | Minimum | Minimum cosine similarity between sentences separated by one intervening sentence |
| soc_mx | Second-order coherence | Maximum | Maximum cosine similarity between sentences separated by one intervening sentence |
| soc_iq | Second-order coherence | Interquartile range | Interquartile range of cosine similarity between sentences separated by one intervening sentence |

**Figure 1-S.** Feature-wide association of semantic similarity metrics and SSD vs HV diagnosis in pre-processing levels 1, 2, and 3; Panels A, B, and C, respectively. Note: glv = GloVe; lsa = LSA; w2v = Word2Vec; rbt = RoBERTa; gpt = GPT-2; lma = Llama.

Panel A

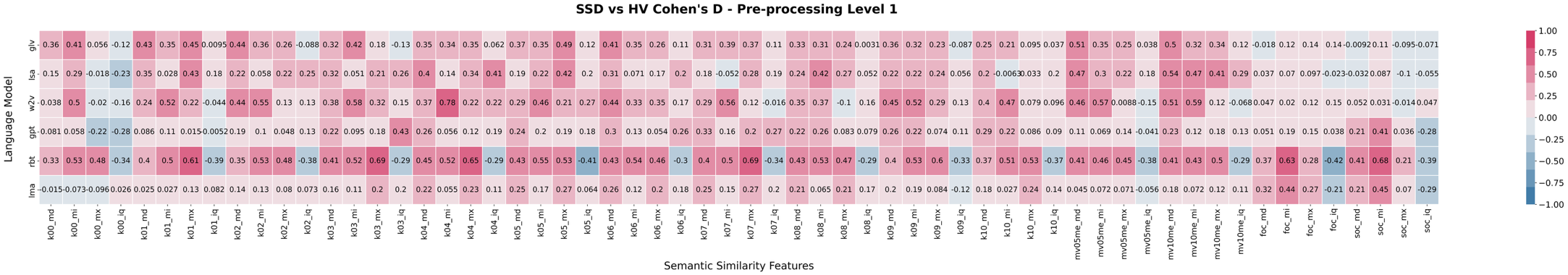

Panel B

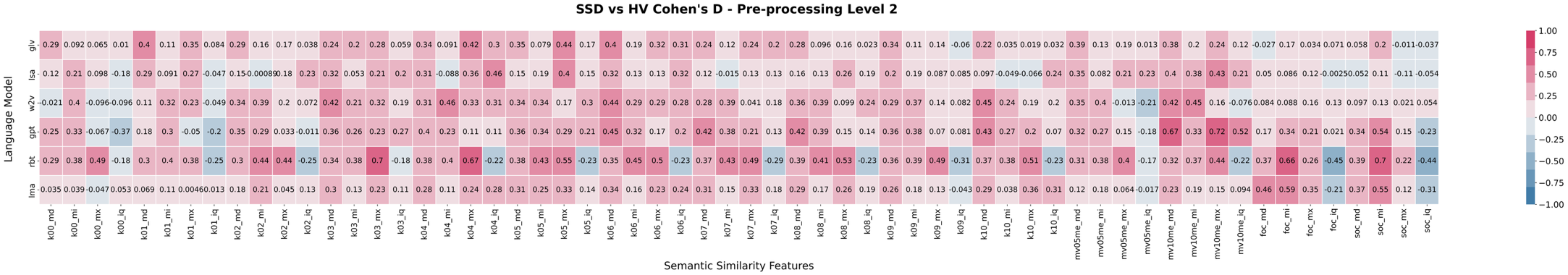

Panel C

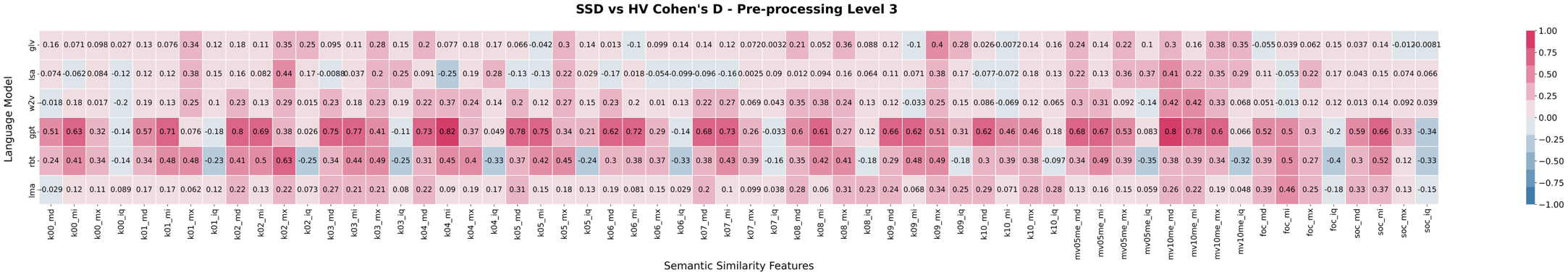

**Table S-2.** Feature-wide statistics of associations between semantic similarity features and SSD diagnosis.

| Pre-processing Level | Language Model | Similarity Feature | Effect Size (d) | Statistical Significance (p) |
| --- | --- | --- | --- | --- |
| 1 | GloVe | k00_md | 0.36 | 0.0195 |
| 1 | GloVe | k00_mi | 0.41 | 0.0114 |
| 1 | GloVe | k00_mx | 0.06 | 0.5588 |
| 1 | GloVe | k00_iq | -0.12 | 0.3438 |
| 1 | GloVe | k01_md | 0.43 | 0.0050 |
| 1 | GloVe | k01_mi | 0.35 | 0.0268 |
| 1 | GloVe | k01_mx | 0.45 | 0.0019 |
| 1 | GloVe | k01_iq | 0.01 | 0.9245 |
| 1 | GloVe | k02_md | 0.44 | 0.0049 |
| 1 | GloVe | k02_mi | 0.36 | 0.0731 |
| 1 | GloVe | k02_mx | 0.26 | 0.0487 |
| 1 | GloVe | k02_iq | -0.09 | 0.8655 |
| 1 | GloVe | k03_md | 0.32 | 0.0214 |
| 1 | GloVe | k03_mi | 0.42 | 0.0045 |
| 1 | GloVe | k03_mx | 0.18 | 0.2603 |
| 1 | GloVe | k03_iq | -0.13 | 0.3614 |
| 1 | GloVe | k04_md | 0.35 | 0.0167 |
| 1 | GloVe | k04_mi | 0.34 | 0.0229 |
| 1 | GloVe | k04_mx | 0.35 | 0.0144 |
| 1 | GloVe | k04_iq | 0.06 | 0.7942 |
| 1 | GloVe | k05_md | 0.37 | 0.0027 |
| 1 | GloVe | k05_mi | 0.35 | 0.0494 |
| 1 | GloVe | k05_mx | 0.49 | 0.0008 |
| 1 | GloVe | k05_iq | 0.12 | 0.2334 |
| 1 | GloVe | k06_md | 0.41 | 0.0041 |
| 1 | GloVe | k06_mi | 0.35 | 0.0584 |
| 1 | GloVe | k06_mx | 0.26 | 0.0762 |
| 1 | GloVe | k06_iq | 0.11 | 0.4363 |
| 1 | GloVe | k07_md | 0.31 | 0.0208 |
| 1 | GloVe | k07_mi | 0.39 | 0.0122 |
| 1 | GloVe | k07_mx | 0.37 | 0.0123 |
| 1 | GloVe | k07_iq | 0.11 | 0.4044 |
| 1 | GloVe | k08_md | 0.33 | 0.0164 |
| 1 | GloVe | k08_mi | 0.31 | 0.0935 |
| 1 | GloVe | k08_mx | 0.24 | 0.1518 |
| 1 | GloVe | k08_iq | 0.00 | 0.7070 |
| 1 | GloVe | k09_md | 0.36 | 0.0079 |
| 1 | GloVe | k09_mi | 0.32 | 0.0114 |
| 1 | GloVe | k09_mx | 0.23 | 0.0565 |
| 1 | GloVe | k09_iq | -0.09 | 0.8840 |
| 1 | GloVe | k10_md | 0.25 | 0.0240 |
| 1 | GloVe | k10_mi | 0.21 | 0.1335 |
| 1 | GloVe | k10_mx | 0.10 | 0.4802 |
| 1 | GloVe | k10_iq | 0.04 | 0.5776 |
| 1 | GloVe | mv05me_md | 0.51 | 0.0017 |
| 1 | GloVe | mv05me_mi | 0.35 | 0.0234 |
| 1 | GloVe | mv05me_mx | 0.25 | 0.1138 |
| 1 | GloVe | mv05me_iq | 0.04 | 0.4935 |
| 1 | GloVe | mv10me_md | 0.50 | 0.0008 |
| 1 | GloVe | mv10me_mi | 0.32 | 0.0408 |
| 1 | GloVe | mv10me_mx | 0.34 | 0.0769 |
| 1 | GloVe | mv10me_iq | 0.12 | 0.6709 |
| 1 | GloVe | foc_md | -0.02 | 0.8901 |
| 1 | GloVe | foc_mi | 0.12 | 0.2226 |
| 1 | GloVe | foc_mx | 0.14 | 0.1688 |
| 1 | GloVe | foc_iq | 0.14 | 0.7038 |
| 1 | GloVe | soc_md | -0.01 | 0.8656 |
| 1 | GloVe | soc_mi | 0.11 | 0.2180 |
| 1 | GloVe | soc_mx | -0.09 | 0.5944 |
| 1 | GloVe | soc_iq | -0.07 | 0.1739 |
| 1 | LSA | k00_md | 0.15 | 0.3570 |
| 1 | LSA | k00_mi | 0.29 | 0.0209 |
| 1 | LSA | k00_mx | -0.02 | 0.8924 |
| 1 | LSA | k00_iq | -0.23 | 0.3173 |
| 1 | LSA | k01_md | 0.35 | 0.0004 |
| 1 | LSA | k01_mi | 0.03 | 0.5660 |
| 1 | LSA | k01_mx | 0.43 | 0.0020 |
| 1 | LSA | k01_iq | 0.18 | 0.0006 |
| 1 | LSA | k02_md | 0.22 | 0.1478 |
| 1 | LSA | k02_mi | 0.06 | 0.3081 |
| 1 | LSA | k02_mx | 0.22 | 0.1828 |
| 1 | LSA | k02_iq | 0.25 | 0.0832 |
| 1 | LSA | k03_md | 0.32 | 0.0091 |
| 1 | LSA | k03_mi | 0.05 | 0.4008 |
| 1 | LSA | k03_mx | 0.21 | 0.1735 |
| 1 | LSA | k03_iq | 0.26 | 0.0610 |
| 1 | LSA | k04_md | 0.40 | 0.0118 |
| 1 | LSA | k04_mi | 0.14 | 0.1672 |
| 1 | LSA | k04_mx | 0.34 | 0.0124 |
| 1 | LSA | k04_iq | 0.41 | 0.0122 |
| 1 | LSA | k05_md | 0.19 | 0.0005 |
| 1 | LSA | k05_mi | 0.22 | 0.1779 |
| 1 | LSA | k05_mx | 0.42 | 0.0027 |
| 1 | LSA | k05_iq | 0.20 | 0.0171 |
| 1 | LSA | k06_md | 0.31 | 0.0163 |
| 1 | LSA | k06_mi | 0.07 | 0.5588 |
| 1 | LSA | k06_mx | 0.17 | 0.1779 |
| 1 | LSA | k06_iq | 0.20 | 0.0923 |
| 1 | LSA | k07_md | 0.18 | 0.4226 |
| 1 | LSA | k07_mi | -0.05 | 0.5878 |
| 1 | LSA | k07_mx | 0.28 | 0.1388 |
| 1 | LSA | k07_iq | 0.19 | 0.0731 |
| 1 | LSA | k08_md | 0.24 | 0.0787 |
| 1 | LSA | k08_mi | 0.42 | 0.0047 |
| 1 | LSA | k08_mx | 0.27 | 0.0599 |
| 1 | LSA | k08_iq | 0.05 | 0.7486 |
| 1 | LSA | k09_md | 0.22 | 0.2070 |
| 1 | LSA | k09_mi | 0.22 | 0.4683 |
| 1 | LSA | k09_mx | 0.24 | 0.0661 |
| 1 | LSA | k09_iq | 0.06 | 0.6834 |
| 1 | LSA | k10_md | 0.20 | 0.1223 |
| 1 | LSA | k10_mi | -0.01 | 0.8146 |
| 1 | LSA | k10_mx | 0.03 | 0.8783 |
| 1 | LSA | k10_iq | 0.20 | 0.2350 |
| 1 | LSA | mv05me_md | 0.47 | 0.0015 |
| 1 | LSA | mv05me_mi | 0.30 | 0.0254 |
| 1 | LSA | mv05me_mx | 0.22 | 0.0935 |
| 1 | LSA | mv05me_iq | 0.18 | 0.1987 |
| 1 | LSA | mv10me_md | 0.54 | 0.0003 |
| 1 | LSA | mv10me_mi | 0.47 | 0.0003 |
| 1 | LSA | mv10me_mx | 0.41 | 0.0034 |
| 1 | LSA | mv10me_iq | 0.29 | 0.0093 |
| 1 | LSA | foc_md | 0.04 | 0.6896 |
| 1 | LSA | foc_mi | 0.07 | 0.2985 |
| 1 | LSA | foc_mx | 0.10 | 0.3805 |
| 1 | LSA | foc_iq | -0.02 | 0.7472 |
| 1 | LSA | soc_md | -0.03 | 0.7446 |
| 1 | LSA | soc_mi | 0.09 | 0.4112 |
| 1 | LSA | soc_mx | -0.10 | 0.2104 |
| 1 | LSA | soc_iq | -0.05 | 0.5608 |
| 1 | Word2Vec | k00_md | 0.04 | 0.1906 |
| 1 | Word2Vec | k00_mi | 0.50 | 0.0002 |
| 1 | Word2Vec | k00_mx | -0.02 | 0.7925 |
| 1 | Word2Vec | k00_iq | -0.16 | 0.8322 |
| 1 | Word2Vec | k01_md | 0.24 | 0.0164 |
| 1 | Word2Vec | k01_mi | 0.52 | 0.0000 |
| 1 | Word2Vec | k01_mx | 0.22 | 0.0813 |
| 1 | Word2Vec | k01_iq | -0.04 | 0.6493 |
| 1 | Word2Vec | k02_md | 0.44 | 0.0032 |
| 1 | Word2Vec | k02_mi | 0.55 | 0.0001 |
| 1 | Word2Vec | k02_mx | 0.13 | 0.3449 |
| 1 | Word2Vec | k02_iq | 0.13 | 0.2505 |
| 1 | Word2Vec | k03_md | 0.38 | 0.0071 |
| 1 | Word2Vec | k03_mi | 0.58 | 0.0000 |
| 1 | Word2Vec | k03_mx | 0.32 | 0.0333 |
| 1 | Word2Vec | k03_iq | 0.15 | 0.1725 |
| 1 | Word2Vec | k04_md | 0.37 | 0.0101 |
| 1 | Word2Vec | k04_mi | 0.78 | 0.0000 |
| 1 | Word2Vec | k04_mx | 0.22 | 0.0956 |
| 1 | Word2Vec | k04_iq | 0.22 | 0.0255 |
| 1 | Word2Vec | k05_md | 0.29 | 0.0041 |
| 1 | Word2Vec | k05_mi | 0.46 | 0.0014 |
| 1 | Word2Vec | k05_mx | 0.21 | 0.1490 |
| 1 | Word2Vec | k05_iq | 0.27 | 0.0343 |
| 1 | Word2Vec | k06_md | 0.44 | 0.0013 |
| 1 | Word2Vec | k06_mi | 0.33 | 0.0042 |
| 1 | Word2Vec | k06_mx | 0.35 | 0.0193 |
| 1 | Word2Vec | k06_iq | 0.17 | 0.1254 |
| 1 | Word2Vec | k07_md | 0.29 | 0.0190 |
| 1 | Word2Vec | k07_mi | 0.56 | 0.0000 |
| 1 | Word2Vec | k07_mx | 0.12 | 0.6655 |
| 1 | Word2Vec | k07_iq | -0.02 | 0.5703 |
| 1 | Word2Vec | k08_md | 0.35 | 0.0050 |
| 1 | Word2Vec | k08_mi | 0.37 | 0.0122 |
| 1 | Word2Vec | k08_mx | -0.10 | 0.3416 |
| 1 | Word2Vec | k08_iq | 0.16 | 0.1964 |
| 1 | Word2Vec | k09_md | 0.45 | 0.0014 |
| 1 | Word2Vec | k09_mi | 0.52 | 0.0000 |
| 1 | Word2Vec | k09_mx | 0.29 | 0.0431 |
| 1 | Word2Vec | k09_iq | 0.13 | 0.2351 |
| 1 | Word2Vec | k10_md | 0.40 | 0.0106 |
| 1 | Word2Vec | k10_mi | 0.47 | 0.0002 |
| 1 | Word2Vec | k10_mx | 0.08 | 0.5573 |
| 1 | Word2Vec | k10_iq | 0.10 | 0.7199 |
| 1 | Word2Vec | mv05me_md | 0.46 | 0.0002 |
| 1 | Word2Vec | mv05me_mi | 0.57 | 0.0003 |
| 1 | Word2Vec | mv05me_mx | 0.01 | 0.6694 |
| 1 | Word2Vec | mv05me_iq | -0.15 | 0.5291 |
| 1 | Word2Vec | mv10me_md | 0.51 | 0.0001 |
| 1 | Word2Vec | mv10me_mi | 0.59 | 0.0000 |
| 1 | Word2Vec | mv10me_mx | 0.12 | 0.2523 |
| 1 | Word2Vec | mv10me_iq | -0.07 | 0.9975 |
| 1 | Word2Vec | foc_md | 0.05 | 0.8682 |
| 1 | Word2Vec | foc_mi | 0.02 | 0.6676 |
| 1 | Word2Vec | foc_mx | 0.12 | 0.2622 |
| 1 | Word2Vec | foc_iq | 0.15 | 0.3455 |
| 1 | Word2Vec | soc_md | 0.05 | 0.7893 |
| 1 | Word2Vec | soc_mi | 0.03 | 0.6725 |
| 1 | Word2Vec | soc_mx | -0.01 | 0.6626 |
| 1 | Word2Vec | soc_iq | 0.05 | 0.7781 |
| 1 | RoBERTa | k00_md | 0.33 | 0.0109 |
| 1 | RoBERTa | k00_mi | 0.53 | 0.0001 |
| 1 | RoBERTa | k00_mx | 0.48 | 0.0002 |
| 1 | RoBERTa | k00_iq | -0.34 | 0.0107 |
| 1 | RoBERTa | k01_md | 0.40 | 0.0030 |
| 1 | RoBERTa | k01_mi | 0.50 | 0.0003 |
| 1 | RoBERTa | k01_mx | 0.61 | 0.0000 |
| 1 | RoBERTa | k01_iq | -0.39 | 0.0060 |
| 1 | RoBERTa | k02_md | 0.35 | 0.0063 |
| 1 | RoBERTa | k02_mi | 0.53 | 0.0002 |
| 1 | RoBERTa | k02_mx | 0.48 | 0.0006 |
| 1 | RoBERTa | k02_iq | -0.38 | 0.0050 |
| 1 | RoBERTa | k03_md | 0.41 | 0.0020 |
| 1 | RoBERTa | k03_mi | 0.52 | 0.0003 |
| 1 | RoBERTa | k03_mx | 0.69 | 0.0000 |
| 1 | RoBERTa | k03_iq | -0.29 | 0.0293 |
| 1 | RoBERTa | k04_md | 0.45 | 0.0007 |
| 1 | RoBERTa | k04_mi | 0.52 | 0.0004 |
| 1 | RoBERTa | k04_mx | 0.65 | 0.0000 |
| 1 | RoBERTa | k04_iq | -0.29 | 0.0438 |
| 1 | RoBERTa | k05_md | 0.43 | 0.0010 |
| 1 | RoBERTa | k05_mi | 0.55 | 0.0001 |
| 1 | RoBERTa | k05_mx | 0.53 | 0.0001 |
| 1 | RoBERTa | k05_iq | -0.41 | 0.0027 |
| 1 | RoBERTa | k06_md | 0.43 | 0.0018 |
| 1 | RoBERTa | k06_mi | 0.54 | 0.0002 |
| 1 | RoBERTa | k06_mx | 0.46 | 0.0006 |
| 1 | RoBERTa | k06_iq | -0.30 | 0.0425 |
| 1 | RoBERTa | k07_md | 0.40 | 0.0039 |
| 1 | RoBERTa | k07_mi | 0.50 | 0.0003 |
| 1 | RoBERTa | k07_mx | 0.69 | 0.0000 |
| 1 | RoBERTa | k07_iq | -0.34 | 0.0155 |
| 1 | RoBERTa | k08_md | 0.43 | 0.0012 |
| 1 | RoBERTa | k08_mi | 0.53 | 0.0002 |
| 1 | RoBERTa | k08_mx | 0.47 | 0.0009 |
| 1 | RoBERTa | k08_iq | -0.29 | 0.0274 |
| 1 | RoBERTa | k09_md | 0.40 | 0.0026 |
| 1 | RoBERTa | k09_mi | 0.53 | 0.0002 |
| 1 | RoBERTa | k09_mx | 0.60 | 0.0000 |
| 1 | RoBERTa | k09_iq | -0.33 | 0.0131 |
| 1 | RoBERTa | k10_md | 0.37 | 0.0025 |
| 1 | RoBERTa | k10_mi | 0.51 | 0.0004 |
| 1 | RoBERTa | k10_mx | 0.53 | 0.0001 |
| 1 | RoBERTa | k10_iq | -0.37 | 0.0090 |
| 1 | RoBERTa | mv05me_md | 0.41 | 0.0022 |
| 1 | RoBERTa | mv05me_mi | 0.46 | 0.0014 |
| 1 | RoBERTa | mv05me_mx | 0.45 | 0.0005 |
| 1 | RoBERTa | mv05me_iq | -0.38 | 0.0148 |
| 1 | RoBERTa | mv10me_md | 0.41 | 0.0022 |
| 1 | RoBERTa | mv10me_mi | 0.43 | 0.0021 |
| 1 | RoBERTa | mv10me_mx | 0.50 | 0.0002 |
| 1 | RoBERTa | mv10me_iq | -0.29 | 0.0258 |
| 1 | RoBERTa | foc_md | 0.37 | 0.0006 |
| 1 | RoBERTa | foc_mi | 0.63 | 0.0000 |
| 1 | RoBERTa | foc_mx | 0.28 | 0.0046 |
| 1 | RoBERTa | foc_iq | -0.42 | 0.0017 |
| 1 | RoBERTa | soc_md | 0.41 | 0.0105 |
| 1 | RoBERTa | soc_mi | 0.68 | 0.0000 |
| 1 | RoBERTa | soc_mx | 0.21 | 0.8369 |
| 1 | RoBERTa | soc_iq | -0.39 | 0.0031 |
| 1 | GPT-2 | k00_md | 0.08 | 0.4749 |
| 1 | GPT-2 | k00_mi | 0.06 | 0.8007 |
| 1 | GPT-2 | k00_mx | -0.22 | 0.1324 |
| 1 | GPT-2 | k00_iq | -0.28 | 0.0254 |
| 1 | GPT-2 | k01_md | 0.09 | 0.4528 |
| 1 | GPT-2 | k01_mi | 0.11 | 0.5996 |
| 1 | GPT-2 | k01_mx | 0.02 | 0.6570 |
| 1 | GPT-2 | k01_iq | -0.01 | 0.7664 |
| 1 | GPT-2 | k02_md | 0.19 | 0.2479 |
| 1 | GPT-2 | k02_mi | 0.10 | 0.2807 |
| 1 | GPT-2 | k02_mx | 0.05 | 0.8622 |
| 1 | GPT-2 | k02_iq | 0.13 | 0.2952 |
| 1 | GPT-2 | k03_md | 0.22 | 0.1484 |
| 1 | GPT-2 | k03_mi | 0.09 | 0.6040 |
| 1 | GPT-2 | k03_mx | 0.18 | 0.3925 |
| 1 | GPT-2 | k03_iq | 0.43 | 0.0057 |
| 1 | GPT-2 | k04_md | 0.26 | 0.1414 |
| 1 | GPT-2 | k04_mi | 0.06 | 0.6585 |
| 1 | GPT-2 | k04_mx | 0.12 | 0.3866 |
| 1 | GPT-2 | k04_iq | 0.19 | 0.2640 |
| 1 | GPT-2 | k05_md | 0.24 | 0.1259 |
| 1 | GPT-2 | k05_mi | 0.20 | 0.2649 |
| 1 | GPT-2 | k05_mx | 0.19 | 0.0769 |
| 1 | GPT-2 | k05_iq | 0.18 | 0.0502 |
| 1 | GPT-2 | k06_md | 0.30 | 0.0381 |
| 1 | GPT-2 | k06_mi | 0.13 | 0.3548 |
| 1 | GPT-2 | k06_mx | 0.05 | 0.5790 |
| 1 | GPT-2 | k06_iq | 0.26 | 0.0576 |
| 1 | GPT-2 | k07_md | 0.33 | 0.0225 |
| 1 | GPT-2 | k07_mi | 0.16 | 0.3011 |
| 1 | GPT-2 | k07_mx | 0.20 | 0.1203 |
| 1 | GPT-2 | k07_iq | 0.27 | 0.0222 |
| 1 | GPT-2 | k08_md | 0.22 | 0.0882 |
| 1 | GPT-2 | k08_mi | 0.26 | 0.0927 |
| 1 | GPT-2 | k08_mx | 0.08 | 0.3112 |
| 1 | GPT-2 | k08_iq | 0.08 | 0.4128 |
| 1 | GPT-2 | k09_md | 0.26 | 0.0668 |
| 1 | GPT-2 | k09_mi | 0.22 | 0.1570 |
| 1 | GPT-2 | k09_mx | 0.07 | 0.3471 |
| 1 | GPT-2 | k09_iq | 0.11 | 0.2385 |
| 1 | GPT-2 | k10_md | 0.29 | 0.0585 |
| 1 | GPT-2 | k10_mi | 0.22 | 0.1109 |
| 1 | GPT-2 | k10_mx | 0.09 | 0.3994 |
| 1 | GPT-2 | k10_iq | 0.09 | 0.4042 |
| 1 | GPT-2 | mv05me_md | 0.11 | 0.4644 |
| 1 | GPT-2 | mv05me_mi | 0.07 | 0.4775 |
| 1 | GPT-2 | mv05me_mx | 0.14 | 0.2972 |
| 1 | GPT-2 | mv05me_iq | -0.04 | 0.3236 |
| 1 | GPT-2 | mv10me_md | 0.23 | 0.1490 |
| 1 | GPT-2 | mv10me_mi | 0.12 | 0.2171 |
| 1 | GPT-2 | mv10me_mx | 0.18 | 0.3041 |
| 1 | GPT-2 | mv10me_iq | 0.13 | 0.6432 |
| 1 | GPT-2 | foc_md | 0.05 | 0.6802 |
| 1 | GPT-2 | foc_mi | 0.19 | 0.1804 |
| 1 | GPT-2 | foc_mx | 0.15 | 0.1889 |
| 1 | GPT-2 | foc_iq | 0.04 | 0.9821 |
| 1 | GPT-2 | soc_md | 0.21 | 0.2516 |
| 1 | GPT-2 | soc_mi | 0.41 | 0.0062 |
| 1 | GPT-2 | soc_mx | 0.04 | 0.6168 |
| 1 | GPT-2 | soc_iq | -0.28 | 0.0249 |
| 1 | Llama | k00_md | -0.02 | 0.9431 |
| 1 | Llama | k00_mi | -0.07 | 0.6040 |
| 1 | Llama | k00_mx | -0.10 | 0.6616 |
| 1 | Llama | k00_iq | 0.03 | 0.9618 |
| 1 | Llama | k01_md | 0.02 | 0.6771 |
| 1 | Llama | k01_mi | 0.03 | 0.8455 |
| 1 | Llama | k01_mx | 0.13 | 0.0787 |
| 1 | Llama | k01_iq | 0.08 | 0.3960 |
| 1 | Llama | k02_md | 0.14 | 0.2393 |
| 1 | Llama | k02_mi | 0.13 | 0.3246 |
| 1 | Llama | k02_mx | 0.08 | 0.4935 |
| 1 | Llama | k02_iq | 0.07 | 0.6756 |
| 1 | Llama | k03_md | 0.16 | 0.3163 |
| 1 | Llama | k03_mi | 0.11 | 0.4350 |
| 1 | Llama | k03_mx | 0.20 | 0.2260 |
| 1 | Llama | k03_iq | 0.20 | 0.1192 |
| 1 | Llama | k04_md | 0.22 | 0.0898 |
| 1 | Llama | k04_mi | 0.05 | 0.5937 |
| 1 | Llama | k04_mx | 0.23 | 0.0404 |
| 1 | Llama | k04_iq | 0.11 | 0.6447 |
| 1 | Llama | k05_md | 0.25 | 0.0886 |
| 1 | Llama | k05_mi | 0.17 | 0.2836 |
| 1 | Llama | k05_mx | 0.27 | 0.0054 |
| 1 | Llama | k05_iq | 0.06 | 0.2962 |
| 1 | Llama | k06_md | 0.26 | 0.0490 |
| 1 | Llama | k06_mi | 0.12 | 0.3536 |
| 1 | Llama | k06_mx | 0.20 | 0.0443 |
| 1 | Llama | k06_iq | 0.18 | 0.2630 |
| 1 | Llama | k07_md | 0.25 | 0.0461 |
| 1 | Llama | k07_mi | 0.15 | 0.3503 |
| 1 | Llama | k07_mx | 0.27 | 0.0144 |
| 1 | Llama | k07_iq | 0.20 | 0.1329 |
| 1 | Llama | k08_md | 0.21 | 0.1259 |
| 1 | Llama | k08_mi | 0.06 | 0.4828 |
| 1 | Llama | k08_mx | 0.21 | 0.0250 |
| 1 | Llama | k08_iq | 0.17 | 0.2826 |
| 1 | Llama | k09_md | 0.20 | 0.1123 |
| 1 | Llama | k09_mi | 0.19 | 0.2704 |
| 1 | Llama | k09_mx | 0.08 | 0.0787 |
| 1 | Llama | k09_iq | -0.12 | 0.6386 |
| 1 | Llama | k10_md | 0.18 | 0.1331 |
| 1 | Llama | k10_mi | 0.03 | 0.8303 |
| 1 | Llama | k10_mx | 0.24 | 0.0314 |
| 1 | Llama | k10_iq | 0.14 | 0.6242 |
| 1 | Llama | mv05me_md | 0.04 | 0.6678 |
| 1 | Llama | mv05me_mi | 0.07 | 0.6663 |
| 1 | Llama | mv05me_mx | 0.07 | 0.4502 |
| 1 | Llama | mv05me_iq | -0.06 | 0.5617 |
| 1 | Llama | mv10me_md | 0.18 | 0.1759 |
| 1 | Llama | mv10me_mi | 0.07 | 0.5291 |
| 1 | Llama | mv10me_mx | 0.12 | 0.2981 |
| 1 | Llama | mv10me_iq | 0.11 | 0.7453 |
| 1 | Llama | foc_md | 0.32 | 0.0378 |
| 1 | Llama | foc_mi | 0.44 | 0.0011 |
| 1 | Llama | foc_mx | 0.27 | 0.0351 |
| 1 | Llama | foc_iq | -0.21 | 0.0333 |
| 1 | Llama | soc_md | 0.21 | 0.1514 |
| 1 | Llama | soc_mi | 0.45 | 0.0022 |
| 1 | Llama | soc_mx | 0.07 | 0.2591 |
| 1 | Llama | soc_iq | -0.29 | 0.0312 |
| 2 | GloVe | k00_md | 0.29 | 0.0894 |
| 2 | GloVe | k00_mi | 0.09 | 0.7534 |
| 2 | GloVe | k00_mx | 0.07 | 0.6092 |
| 2 | GloVe | k00_iq | 0.01 | 0.9652 |
| 2 | GloVe | k01_md | 0.40 | 0.0074 |
| 2 | GloVe | k01_mi | 0.11 | 0.5996 |
| 2 | GloVe | k01_mx | 0.35 | 0.0191 |
| 2 | GloVe | k01_iq | 0.08 | 0.4709 |
| 2 | GloVe | k02_md | 0.29 | 0.0530 |
| 2 | GloVe | k02_mi | 0.16 | 0.3405 |
| 2 | GloVe | k02_mx | 0.17 | 0.1998 |
| 2 | GloVe | k02_iq | 0.04 | 0.1104 |
| 2 | GloVe | k03_md | 0.24 | 0.0368 |
| 2 | GloVe | k03_mi | 0.20 | 0.1455 |
| 2 | GloVe | k03_mx | 0.28 | 0.0802 |
| 2 | GloVe | k03_iq | 0.06 | 0.3081 |
| 2 | GloVe | k04_md | 0.34 | 0.0143 |
| 2 | GloVe | k04_mi | 0.09 | 0.5937 |
| 2 | GloVe | k04_mx | 0.42 | 0.0040 |
| 2 | GloVe | k04_iq | 0.30 | 0.0527 |
| 2 | GloVe | k05_md | 0.35 | 0.0106 |
| 2 | GloVe | k05_mi | 0.08 | 0.8007 |
| 2 | GloVe | k05_mx | 0.44 | 0.0022 |
| 2 | GloVe | k05_iq | 0.17 | 0.1048 |
| 2 | GloVe | k06_md | 0.40 | 0.0087 |
| 2 | GloVe | k06_mi | 0.19 | 0.3320 |
| 2 | GloVe | k06_mx | 0.32 | 0.0161 |
| 2 | GloVe | k06_iq | 0.31 | 0.0209 |
| 2 | GloVe | k07_md | 0.24 | 0.0353 |
| 2 | GloVe | k07_mi | 0.12 | 0.6190 |
| 2 | GloVe | k07_mx | 0.24 | 0.0561 |
| 2 | GloVe | k07_iq | 0.20 | 0.1003 |
| 2 | GloVe | k08_md | 0.28 | 0.0231 |
| 2 | GloVe | k08_mi | 0.10 | 0.5166 |
| 2 | GloVe | k08_mx | 0.16 | 0.2685 |
| 2 | GloVe | k08_iq | 0.02 | 0.7729 |
| 2 | GloVe | k09_md | 0.34 | 0.0148 |
| 2 | GloVe | k09_mi | 0.11 | 0.5732 |
| 2 | GloVe | k09_mx | 0.14 | 0.1679 |
| 2 | GloVe | k09_iq | -0.06 | 0.9363 |
| 2 | GloVe | k10_md | 0.22 | 0.0517 |
| 2 | GloVe | k10_mi | 0.04 | 0.9313 |
| 2 | GloVe | k10_mx | 0.02 | 0.8073 |
| 2 | GloVe | k10_iq | 0.03 | 0.5849 |
| 2 | GloVe | mv05me_md | 0.39 | 0.0135 |
| 2 | GloVe | mv05me_mi | 0.13 | 0.4657 |
| 2 | GloVe | mv05me_mx | 0.19 | 0.2962 |
| 2 | GloVe | mv05me_iq | 0.01 | 0.4881 |
| 2 | GloVe | mv10me_md | 0.38 | 0.0069 |
| 2 | GloVe | mv10me_mi | 0.20 | 0.2101 |
| 2 | GloVe | mv10me_mx | 0.24 | 0.2630 |
| 2 | GloVe | mv10me_iq | 0.12 | 0.4288 |
| 2 | GloVe | foc_md | -0.03 | 0.7407 |
| 2 | GloVe | foc_mi | 0.17 | 0.1330 |
| 2 | GloVe | foc_mx | 0.03 | 0.2985 |
| 2 | GloVe | foc_iq | 0.07 | 0.9547 |
| 2 | GloVe | soc_md | 0.06 | 0.6824 |
| 2 | GloVe | soc_mi | 0.20 | 0.0898 |
| 2 | GloVe | soc_mx | -0.01 | 0.8719 |
| 2 | GloVe | soc_iq | -0.04 | 0.2299 |
| 2 | LSA | k00_md | 0.12 | 0.4496 |
| 2 | LSA | k00_mi | 0.21 | 0.0672 |
| 2 | LSA | k00_mx | 0.10 | 0.6920 |
| 2 | LSA | k00_iq | -0.18 | 0.3031 |
| 2 | LSA | k01_md | 0.29 | 0.0029 |
| 2 | LSA | k01_mi | 0.09 | 0.2195 |
| 2 | LSA | k01_mx | 0.27 | 0.0271 |
| 2 | LSA | k01_iq | -0.05 | 0.0353 |
| 2 | LSA | k02_md | 0.15 | 0.3071 |
| 2 | LSA | k02_mi | 0.00 | 0.6463 |
| 2 | LSA | k02_mx | 0.18 | 0.3733 |
| 2 | LSA | k02_iq | 0.23 | 0.1265 |
| 2 | LSA | k03_md | 0.32 | 0.0143 |
| 2 | LSA | k03_mi | 0.05 | 0.8472 |
| 2 | LSA | k03_mx | 0.21 | 0.0906 |
| 2 | LSA | k03_iq | 0.20 | 0.1814 |
| 2 | LSA | k04_md | 0.31 | 0.1119 |
| 2 | LSA | k04_mi | -0.09 | 0.8975 |
| 2 | LSA | k04_mx | 0.36 | 0.0167 |
| 2 | LSA | k04_iq | 0.46 | 0.0023 |
| 2 | LSA | k05_md | 0.15 | 0.0088 |
| 2 | LSA | k05_mi | 0.19 | 0.3051 |
| 2 | LSA | k05_mx | 0.40 | 0.0036 |
| 2 | LSA | k05_iq | 0.15 | 0.1329 |
| 2 | LSA | k06_md | 0.32 | 0.0137 |
| 2 | LSA | k06_mi | 0.13 | 0.3071 |
| 2 | LSA | k06_mx | 0.13 | 0.3438 |
| 2 | LSA | k06_iq | 0.32 | 0.0133 |
| 2 | LSA | k07_md | 0.12 | 0.6771 |
| 2 | LSA | k07_mi | -0.01 | 0.5893 |
| 2 | LSA | k07_mx | 0.13 | 0.3581 |
| 2 | LSA | k07_iq | 0.13 | 0.6417 |
| 2 | LSA | k08_md | 0.16 | 0.2567 |
| 2 | LSA | k08_mi | 0.13 | 0.7070 |
| 2 | LSA | k08_mx | 0.26 | 0.0258 |
| 2 | LSA | k08_iq | 0.19 | 0.2054 |
| 2 | LSA | k09_md | 0.20 | 0.1877 |
| 2 | LSA | k09_mi | 0.19 | 0.2807 |
| 2 | LSA | k09_mx | 0.09 | 0.3362 |
| 2 | LSA | k09_iq | 0.08 | 0.6740 |
| 2 | LSA | k10_md | 0.10 | 0.5740 |
| 2 | LSA | k10_mi | -0.05 | 0.7553 |
| 2 | LSA | k10_mx | -0.07 | 0.5488 |
| 2 | LSA | k10_iq | 0.24 | 0.1177 |
| 2 | LSA | mv05me_md | 0.35 | 0.0077 |
| 2 | LSA | mv05me_mi | 0.08 | 0.5125 |
| 2 | LSA | mv05me_mx | 0.21 | 0.0582 |
| 2 | LSA | mv05me_iq | 0.23 | 0.1259 |
| 2 | LSA | mv10me_md | 0.40 | 0.0046 |
| 2 | LSA | mv10me_mi | 0.38 | 0.0027 |
| 2 | LSA | mv10me_mx | 0.43 | 0.0027 |
| 2 | LSA | mv10me_iq | 0.21 | 0.0438 |
| 2 | LSA | foc_md | 0.05 | 0.6503 |
| 2 | LSA | foc_mi | 0.09 | 0.2144 |
| 2 | LSA | foc_mx | 0.12 | 0.2831 |
| 2 | LSA | foc_iq | 0.00 | 0.8472 |
| 2 | LSA | soc_md | -0.05 | 0.6080 |
| 2 | LSA | soc_mi | 0.11 | 0.3163 |
| 2 | LSA | soc_mx | -0.11 | 0.2290 |
| 2 | LSA | soc_iq | -0.05 | 0.5845 |
| 2 | Word2Vec | k00_md | -0.02 | 0.3481 |
| 2 | Word2Vec | k00_mi | 0.40 | 0.0049 |
| 2 | Word2Vec | k00_mx | -0.10 | 0.4605 |
| 2 | Word2Vec | k00_iq | -0.10 | 0.8405 |
| 2 | Word2Vec | k01_md | 0.11 | 0.0551 |
| 2 | Word2Vec | k01_mi | 0.32 | 0.0107 |
| 2 | Word2Vec | k01_mx | 0.23 | 0.0497 |
| 2 | Word2Vec | k01_iq | -0.05 | 0.1633 |
| 2 | Word2Vec | k02_md | 0.34 | 0.0108 |
| 2 | Word2Vec | k02_mi | 0.39 | 0.0075 |
| 2 | Word2Vec | k02_mx | 0.20 | 0.1425 |
| 2 | Word2Vec | k02_iq | 0.07 | 0.1340 |
| 2 | Word2Vec | k03_md | 0.42 | 0.0041 |
| 2 | Word2Vec | k03_mi | 0.21 | 0.0787 |
| 2 | Word2Vec | k03_mx | 0.32 | 0.0268 |
| 2 | Word2Vec | k03_iq | 0.19 | 0.0475 |
| 2 | Word2Vec | k04_md | 0.31 | 0.0257 |
| 2 | Word2Vec | k04_mi | 0.46 | 0.0010 |
| 2 | Word2Vec | k04_mx | 0.33 | 0.0129 |
| 2 | Word2Vec | k04_iq | 0.31 | 0.0152 |
| 2 | Word2Vec | k05_md | 0.34 | 0.0015 |
| 2 | Word2Vec | k05_mi | 0.34 | 0.0087 |
| 2 | Word2Vec | k05_mx | 0.17 | 0.3092 |
| 2 | Word2Vec | k05_iq | 0.30 | 0.0089 |
| 2 | Word2Vec | k06_md | 0.44 | 0.0015 |
| 2 | Word2Vec | k06_mi | 0.29 | 0.0286 |
| 2 | Word2Vec | k06_mx | 0.29 | 0.0886 |
| 2 | Word2Vec | k06_iq | 0.28 | 0.0247 |
| 2 | Word2Vec | k07_md | 0.28 | 0.0477 |
| 2 | Word2Vec | k07_mi | 0.39 | 0.0041 |
| 2 | Word2Vec | k07_mx | 0.04 | 0.8664 |
| 2 | Word2Vec | k07_iq | 0.18 | 0.1153 |
| 2 | Word2Vec | k08_md | 0.36 | 0.0080 |
| 2 | Word2Vec | k08_mi | 0.39 | 0.0016 |
| 2 | Word2Vec | k08_mx | 0.10 | 0.5229 |
| 2 | Word2Vec | k08_iq | 0.24 | 0.1039 |
| 2 | Word2Vec | k09_md | 0.29 | 0.0605 |
| 2 | Word2Vec | k09_mi | 0.37 | 0.0026 |
| 2 | Word2Vec | k09_mx | 0.14 | 0.4128 |
| 2 | Word2Vec | k09_iq | 0.08 | 0.4631 |
| 2 | Word2Vec | k10_md | 0.45 | 0.0041 |
| 2 | Word2Vec | k10_mi | 0.24 | 0.0871 |
| 2 | Word2Vec | k10_mx | 0.19 | 0.2860 |
| 2 | Word2Vec | k10_iq | 0.20 | 0.0828 |
| 2 | Word2Vec | mv05me_md | 0.35 | 0.0005 |
| 2 | Word2Vec | mv05me_mi | 0.40 | 0.0164 |
| 2 | Word2Vec | mv05me_mx | -0.01 | 0.5996 |
| 2 | Word2Vec | mv05me_iq | -0.21 | 0.3153 |
| 2 | Word2Vec | mv10me_md | 0.42 | 0.0003 |
| 2 | Word2Vec | mv10me_mi | 0.45 | 0.0007 |
| 2 | Word2Vec | mv10me_mx | 0.16 | 0.1380 |
| 2 | Word2Vec | mv10me_iq | -0.08 | 0.8605 |
| 2 | Word2Vec | foc_md | 0.08 | 0.6457 |
| 2 | Word2Vec | foc_mi | 0.09 | 0.3756 |
| 2 | Word2Vec | foc_mx | 0.16 | 0.2308 |
| 2 | Word2Vec | foc_iq | 0.13 | 0.4250 |
| 2 | Word2Vec | soc_md | 0.10 | 0.7824 |
| 2 | Word2Vec | soc_mi | 0.13 | 0.2334 |
| 2 | Word2Vec | soc_mx | 0.02 | 0.9308 |
| 2 | Word2Vec | soc_iq | 0.05 | 0.7841 |
| 2 | RoBERTa | k00_md | 0.29 | 0.0391 |
| 2 | RoBERTa | k00_mi | 0.38 | 0.0018 |
| 2 | RoBERTa | k00_mx | 0.49 | 0.0005 |
| 2 | RoBERTa | k00_iq | -0.18 | 0.2470 |
| 2 | RoBERTa | k01_md | 0.30 | 0.0192 |
| 2 | RoBERTa | k01_mi | 0.40 | 0.0014 |
| 2 | RoBERTa | k01_mx | 0.38 | 0.0014 |
| 2 | RoBERTa | k01_iq | -0.25 | 0.0538 |
| 2 | RoBERTa | k02_md | 0.30 | 0.0267 |
| 2 | RoBERTa | k02_mi | 0.44 | 0.0011 |
| 2 | RoBERTa | k02_mx | 0.44 | 0.0010 |
| 2 | RoBERTa | k02_iq | -0.25 | 0.0468 |
| 2 | RoBERTa | k03_md | 0.34 | 0.0160 |
| 2 | RoBERTa | k03_mi | 0.38 | 0.0066 |
| 2 | RoBERTa | k03_mx | 0.70 | 0.0000 |
| 2 | RoBERTa | k03_iq | -0.18 | 0.1162 |
| 2 | RoBERTa | k04_md | 0.38 | 0.0051 |
| 2 | RoBERTa | k04_mi | 0.40 | 0.0028 |
| 2 | RoBERTa | k04_mx | 0.67 | 0.0000 |
| 2 | RoBERTa | k04_iq | -0.22 | 0.0999 |
| 2 | RoBERTa | k05_md | 0.38 | 0.0033 |
| 2 | RoBERTa | k05_mi | 0.43 | 0.0012 |
| 2 | RoBERTa | k05_mx | 0.55 | 0.0001 |
| 2 | RoBERTa | k05_iq | -0.23 | 0.0724 |
| 2 | RoBERTa | k06_md | 0.35 | 0.0053 |
| 2 | RoBERTa | k06_mi | 0.45 | 0.0011 |
| 2 | RoBERTa | k06_mx | 0.50 | 0.0004 |
| 2 | RoBERTa | k06_iq | -0.23 | 0.1308 |
| 2 | RoBERTa | k07_md | 0.37 | 0.0078 |
| 2 | RoBERTa | k07_mi | 0.43 | 0.0006 |
| 2 | RoBERTa | k07_mx | 0.49 | 0.0005 |
| 2 | RoBERTa | k07_iq | -0.29 | 0.0227 |
| 2 | RoBERTa | k08_md | 0.39 | 0.0037 |
| 2 | RoBERTa | k08_mi | 0.41 | 0.0019 |
| 2 | RoBERTa | k08_mx | 0.53 | 0.0002 |
| 2 | RoBERTa | k08_iq | -0.23 | 0.0824 |
| 2 | RoBERTa | k09_md | 0.36 | 0.0079 |
| 2 | RoBERTa | k09_mi | 0.39 | 0.0020 |
| 2 | RoBERTa | k09_mx | 0.49 | 0.0003 |
| 2 | RoBERTa | k09_iq | -0.31 | 0.0244 |
| 2 | RoBERTa | k10_md | 0.37 | 0.0078 |
| 2 | RoBERTa | k10_mi | 0.38 | 0.0033 |
| 2 | RoBERTa | k10_mx | 0.51 | 0.0004 |
| 2 | RoBERTa | k10_iq | -0.23 | 0.0391 |
| 2 | RoBERTa | mv05me_md | 0.31 | 0.0197 |
| 2 | RoBERTa | mv05me_mi | 0.38 | 0.0062 |
| 2 | RoBERTa | mv05me_mx | 0.40 | 0.0020 |
| 2 | RoBERTa | mv05me_iq | -0.17 | 0.2488 |
| 2 | RoBERTa | mv10me_md | 0.32 | 0.0157 |
| 2 | RoBERTa | mv10me_mi | 0.37 | 0.0082 |
| 2 | RoBERTa | mv10me_mx | 0.44 | 0.0012 |
| 2 | RoBERTa | mv10me_iq | -0.22 | 0.1640 |
| 2 | RoBERTa | foc_md | 0.37 | 0.0007 |
| 2 | RoBERTa | foc_mi | 0.66 | 0.0000 |
| 2 | RoBERTa | foc_mx | 0.26 | 0.0024 |
| 2 | RoBERTa | foc_iq | -0.45 | 0.0007 |
| 2 | RoBERTa | soc_md | 0.39 | 0.0053 |
| 2 | RoBERTa | soc_mi | 0.70 | 0.0000 |
| 2 | RoBERTa | soc_mx | 0.22 | 0.5470 |
| 2 | RoBERTa | soc_iq | -0.44 | 0.0011 |
| 2 | GPT-2 | k00_md | 0.25 | 0.0714 |
| 2 | GPT-2 | k00_mi | 0.33 | 0.0116 |
| 2 | GPT-2 | k00_mx | -0.07 | 0.5305 |
| 2 | GPT-2 | k00_iq | -0.37 | 0.0071 |
| 2 | GPT-2 | k01_md | 0.18 | 0.1877 |
| 2 | GPT-2 | k01_mi | 0.30 | 0.0280 |
| 2 | GPT-2 | k01_mx | -0.05 | 0.9754 |
| 2 | GPT-2 | k01_iq | -0.20 | 0.2219 |
| 2 | GPT-2 | k02_md | 0.35 | 0.0246 |
| 2 | GPT-2 | k02_mi | 0.29 | 0.0490 |
| 2 | GPT-2 | k02_mx | 0.03 | 0.7975 |
| 2 | GPT-2 | k02_iq | -0.01 | 0.8206 |
| 2 | GPT-2 | k03_md | 0.36 | 0.0197 |
| 2 | GPT-2 | k03_mi | 0.26 | 0.1035 |
| 2 | GPT-2 | k03_mx | 0.23 | 0.2252 |
| 2 | GPT-2 | k03_iq | 0.27 | 0.0766 |
| 2 | GPT-2 | k04_md | 0.40 | 0.0134 |
| 2 | GPT-2 | k04_mi | 0.23 | 0.1057 |
| 2 | GPT-2 | k04_mx | 0.11 | 0.4451 |
| 2 | GPT-2 | k04_iq | 0.11 | 0.6447 |
| 2 | GPT-2 | k05_md | 0.36 | 0.0156 |
| 2 | GPT-2 | k05_mi | 0.34 | 0.0347 |
| 2 | GPT-2 | k05_mx | 0.29 | 0.0106 |
| 2 | GPT-2 | k05_iq | 0.21 | 0.1502 |
| 2 | GPT-2 | k06_md | 0.45 | 0.0033 |
| 2 | GPT-2 | k06_mi | 0.32 | 0.0395 |
| 2 | GPT-2 | k06_mx | 0.17 | 0.2845 |
| 2 | GPT-2 | k06_iq | 0.20 | 0.1044 |
| 2 | GPT-2 | k07_md | 0.42 | 0.0048 |
| 2 | GPT-2 | k07_mi | 0.38 | 0.0122 |
| 2 | GPT-2 | k07_mx | 0.21 | 0.0678 |
| 2 | GPT-2 | k07_iq | 0.13 | 0.2317 |
| 2 | GPT-2 | k08_md | 0.42 | 0.0071 |
| 2 | GPT-2 | k08_mi | 0.39 | 0.0099 |
| 2 | GPT-2 | k08_mx | 0.15 | 0.1478 |
| 2 | GPT-2 | k08_iq | 0.14 | 0.2062 |
| 2 | GPT-2 | k09_md | 0.36 | 0.0187 |
| 2 | GPT-2 | k09_mi | 0.38 | 0.0124 |
| 2 | GPT-2 | k09_mx | 0.07 | 0.3492 |
| 2 | GPT-2 | k09_iq | 0.08 | 0.2260 |
| 2 | GPT-2 | k10_md | 0.43 | 0.0058 |
| 2 | GPT-2 | k10_mi | 0.27 | 0.0889 |
| 2 | GPT-2 | k10_mx | 0.20 | 0.0901 |
| 2 | GPT-2 | k10_iq | 0.07 | 0.7832 |
| 2 | GPT-2 | mv05me_md | 0.32 | 0.0492 |
| 2 | GPT-2 | mv05me_mi | 0.27 | 0.0307 |
| 2 | GPT-2 | mv05me_mx | 0.15 | 0.3728 |
| 2 | GPT-2 | mv05me_iq | -0.18 | 0.0263 |
| 2 | GPT-2 | mv10me_md | 0.67 | 0.0000 |
| 2 | GPT-2 | mv10me_mi | 0.33 | 0.0150 |
| 2 | GPT-2 | mv10me_mx | 0.72 | 0.0000 |
| 2 | GPT-2 | mv10me_iq | 0.52 | 0.0002 |
| 2 | GPT-2 | foc_md | 0.17 | 0.2541 |
| 2 | GPT-2 | foc_mi | 0.34 | 0.0230 |
| 2 | GPT-2 | foc_mx | 0.21 | 0.1080 |
| 2 | GPT-2 | foc_iq | 0.02 | 0.9735 |
| 2 | GPT-2 | soc_md | 0.34 | 0.0314 |
| 2 | GPT-2 | soc_mi | 0.54 | 0.0005 |
| 2 | GPT-2 | soc_mx | 0.15 | 0.3282 |
| 2 | GPT-2 | soc_iq | -0.23 | 0.0983 |
| 2 | Llama | k00_md | 0.04 | 0.7942 |
| 2 | Llama | k00_mi | 0.04 | 0.8655 |
| 2 | Llama | k00_mx | -0.05 | 0.9805 |
| 2 | Llama | k00_iq | 0.05 | 0.5084 |
| 2 | Llama | k01_md | 0.07 | 0.4439 |
| 2 | Llama | k01_mi | 0.11 | 0.4068 |
| 2 | Llama | k01_mx | 0.00 | 0.3682 |
| 2 | Llama | k01_iq | 0.01 | 0.7892 |
| 2 | Llama | k02_md | 0.18 | 0.2942 |
| 2 | Llama | k02_mi | 0.21 | 0.2497 |
| 2 | Llama | k02_mx | 0.05 | 0.6386 |
| 2 | Llama | k02_iq | 0.13 | 0.3256 |
| 2 | Llama | k03_md | 0.30 | 0.0579 |
| 2 | Llama | k03_mi | 0.13 | 0.3525 |
| 2 | Llama | k03_mx | 0.23 | 0.1570 |
| 2 | Llama | k03_iq | 0.11 | 0.1957 |
| 2 | Llama | k04_md | 0.28 | 0.0659 |
| 2 | Llama | k04_mi | 0.11 | 0.3416 |
| 2 | Llama | k04_mx | 0.24 | 0.0512 |
| 2 | Llama | k04_iq | 0.28 | 0.0593 |
| 2 | Llama | k05_md | 0.31 | 0.0459 |
| 2 | Llama | k05_mi | 0.25 | 0.1685 |
| 2 | Llama | k05_mx | 0.33 | 0.0020 |
| 2 | Llama | k05_iq | 0.14 | 0.1484 |
| 2 | Llama | k06_md | 0.34 | 0.0326 |
| 2 | Llama | k06_mi | 0.16 | 0.3438 |
| 2 | Llama | k06_mx | 0.23 | 0.1026 |
| 2 | Llama | k06_iq | 0.24 | 0.1148 |
| 2 | Llama | k07_md | 0.31 | 0.0247 |
| 2 | Llama | k07_mi | 0.15 | 0.3256 |
| 2 | Llama | k07_mx | 0.33 | 0.0066 |
| 2 | Llama | k07_iq | 0.18 | 0.1807 |
| 2 | Llama | k08_md | 0.29 | 0.0535 |
| 2 | Llama | k08_mi | 0.17 | 0.4502 |
| 2 | Llama | k08_mx | 0.26 | 0.0335 |
| 2 | Llama | k08_iq | 0.19 | 0.1157 |
| 2 | Llama | k09_md | 0.26 | 0.0701 |
| 2 | Llama | k09_mi | 0.18 | 0.2488 |
| 2 | Llama | k09_mx | 0.13 | 0.1286 |
| 2 | Llama | k09_iq | -0.04 | 0.6896 |
| 2 | Llama | k10_md | 0.29 | 0.0720 |
| 2 | Llama | k10_mi | 0.04 | 0.9665 |
| 2 | Llama | k10_mx | 0.36 | 0.0047 |
| 2 | Llama | k10_iq | 0.31 | 0.0272 |
| 2 | Llama | mv05me_md | 0.12 | 0.4128 |
| 2 | Llama | mv05me_mi | 0.18 | 0.2017 |
| 2 | Llama | mv05me_mx | 0.06 | 0.5893 |
| 2 | Llama | mv05me_iq | -0.02 | 0.4775 |
| 2 | Llama | mv10me_md | 0.23 | 0.1533 |
| 2 | Llama | mv10me_mi | 0.19 | 0.1208 |
| 2 | Llama | mv10me_mx | 0.15 | 0.3267 |
| 2 | Llama | mv10me_iq | 0.09 | 0.9059 |
| 2 | Llama | foc_md | 0.46 | 0.0054 |
| 2 | Llama | foc_mi | 0.59 | 0.0000 |
| 2 | Llama | foc_mx | 0.35 | 0.0258 |
| 2 | Llama | foc_iq | -0.21 | 0.0636 |
| 2 | Llama | soc_md | 0.37 | 0.0192 |
| 2 | Llama | soc_mi | 0.55 | 0.0001 |
| 2 | Llama | soc_mx | 0.12 | 0.2713 |
| 2 | Llama | soc_iq | -0.31 | 0.0277 |
| 3 | GloVe | k00_md | 0.16 | 0.2826 |
| 3 | GloVe | k00_mi | 0.07 | 0.8790 |
| 3 | GloVe | k00_mx | 0.10 | 0.4736 |
| 3 | GloVe | k00_iq | 0.03 | 0.5574 |
| 3 | GloVe | k01_md | 0.13 | 0.5488 |
| 3 | GloVe | k01_mi | 0.08 | 0.9346 |
| 3 | GloVe | k01_mx | 0.34 | 0.0299 |
| 3 | GloVe | k01_iq | 0.12 | 0.2101 |
| 3 | GloVe | k02_md | 0.18 | 0.2293 |
| 3 | GloVe | k02_mi | 0.11 | 0.7502 |
| 3 | GloVe | k02_mx | 0.35 | 0.0160 |
| 3 | GloVe | k02_iq | 0.25 | 0.1849 |
| 3 | GloVe | k03_md | 0.09 | 0.4975 |
| 3 | GloVe | k03_mi | 0.11 | 0.8338 |
| 3 | GloVe | k03_mx | 0.28 | 0.0520 |
| 3 | GloVe | k03_iq | 0.15 | 0.2444 |
| 3 | GloVe | k04_md | 0.20 | 0.0840 |
| 3 | GloVe | k04_mi | 0.08 | 0.7737 |
| 3 | GloVe | k04_mx | 0.18 | 0.2707 |
| 3 | GloVe | k04_iq | 0.17 | 0.2393 |
| 3 | GloVe | k05_md | 0.07 | 0.6943 |
| 3 | GloVe | k05_mi | -0.04 | 0.5473 |
| 3 | GloVe | k05_mx | 0.30 | 0.0364 |
| 3 | GloVe | k05_iq | 0.14 | 0.2419 |
| 3 | GloVe | k06_md | 0.01 | 0.7986 |
| 3 | GloVe | k06_mi | -0.10 | 0.5092 |
| 3 | GloVe | k06_mx | 0.10 | 0.5933 |
| 3 | GloVe | k06_iq | 0.14 | 0.3320 |
| 3 | GloVe | k07_md | 0.14 | 0.2968 |
| 3 | GloVe | k07_mi | 0.12 | 0.9237 |
| 3 | GloVe | k07_mx | 0.07 | 0.3151 |
| 3 | GloVe | k07_iq | 0.00 | 0.4414 |
| 3 | GloVe | k08_md | 0.21 | 0.2838 |
| 3 | GloVe | k08_mi | 0.05 | 0.6782 |
| 3 | GloVe | k08_mx | 0.36 | 0.0230 |
| 3 | GloVe | k08_iq | 0.09 | 0.4938 |
| 3 | GloVe | k09_md | 0.12 | 0.2498 |
| 3 | GloVe | k09_mi | -0.10 | 0.1990 |
| 3 | GloVe | k09_mx | 0.40 | 0.0015 |
| 3 | GloVe | k09_iq | 0.28 | 0.0132 |
| 3 | GloVe | k10_md | 0.03 | 0.3901 |
| 3 | GloVe | k10_mi | -0.01 | 0.7926 |
| 3 | GloVe | k10_mx | 0.14 | 0.3288 |
| 3 | GloVe | k10_iq | 0.16 | 0.1778 |
| 3 | GloVe | mv05me_md | 0.24 | 0.1705 |
| 3 | GloVe | mv05me_mi | 0.14 | 0.6709 |
| 3 | GloVe | mv05me_mx | 0.22 | 0.1964 |
| 3 | GloVe | mv05me_iq | 0.10 | 0.2140 |
| 3 | GloVe | mv10me_md | 0.30 | 0.0569 |
| 3 | GloVe | mv10me_mi | 0.16 | 0.3250 |
| 3 | GloVe | mv10me_mx | 0.38 | 0.0117 |
| 3 | GloVe | mv10me_iq | 0.35 | 0.0372 |
| 3 | GloVe | foc_md | -0.05 | 0.7770 |
| 3 | GloVe | foc_mi | 0.04 | 0.4793 |
| 3 | GloVe | foc_mx | 0.06 | 0.3849 |
| 3 | GloVe | foc_iq | 0.15 | 0.2471 |
| 3 | GloVe | soc_md | 0.04 | 0.9178 |
| 3 | GloVe | soc_mi | 0.14 | 0.5028 |
| 3 | GloVe | soc_mx | -0.01 | 0.8480 |
| 3 | GloVe | soc_iq | -0.01 | 0.7812 |
| 3 | LSA | k00_md | 0.07 | 0.3173 |
| 3 | LSA | k00_mi | -0.06 | 0.3438 |
| 3 | LSA | k00_mx | 0.08 | 0.8823 |
| 3 | LSA | k00_iq | -0.12 | 0.8172 |
| 3 | LSA | k01_md | 0.12 | 0.1203 |
| 3 | LSA | k01_mi | 0.12 | 0.5016 |
| 3 | LSA | k01_mx | 0.38 | 0.0136 |
| 3 | LSA | k01_iq | 0.15 | 0.1044 |
| 3 | LSA | k02_md | 0.16 | 0.4515 |
| 3 | LSA | k02_mi | 0.08 | 0.6295 |
| 3 | LSA | k02_mx | 0.44 | 0.0139 |
| 3 | LSA | k02_iq | 0.17 | 0.1340 |
| 3 | LSA | k03_md | -0.01 | 0.6460 |
| 3 | LSA | k03_mi | 0.04 | 0.8179 |
| 3 | LSA | k03_mx | 0.20 | 0.5653 |
| 3 | LSA | k03_iq | 0.25 | 0.0976 |
| 3 | LSA | k04_md | 0.09 | 0.7557 |
| 3 | LSA | k04_mi | -0.25 | 0.3439 |
| 3 | LSA | k04_mx | 0.19 | 0.5748 |
| 3 | LSA | k04_iq | 0.28 | 0.2043 |
| 3 | LSA | k05_md | -0.13 | 0.6181 |
| 3 | LSA | k05_mi | -0.13 | 0.2968 |
| 3 | LSA | k05_mx | 0.22 | 0.7639 |
| 3 | LSA | k05_iq | 0.03 | 0.2074 |
| 3 | LSA | k06_md | -0.17 | 0.7576 |
| 3 | LSA | k06_mi | 0.02 | 0.8143 |
| 3 | LSA | k06_mx | -0.05 | 0.4008 |
| 3 | LSA | k06_iq | -0.10 | 0.8412 |
| 3 | LSA | k07_md | -0.10 | 0.6173 |
| 3 | LSA | k07_mi | -0.16 | 0.9852 |
| 3 | LSA | k07_mx | 0.00 | 0.4389 |
| 3 | LSA | k07_iq | 0.09 | 0.7888 |
| 3 | LSA | k08_md | 0.01 | 0.9736 |
| 3 | LSA | k08_mi | 0.09 | 0.8251 |
| 3 | LSA | k08_mx | 0.16 | 0.4906 |
| 3 | LSA | k08_iq | 0.06 | 0.3999 |
| 3 | LSA | k09_md | 0.11 | 0.2008 |
| 3 | LSA | k09_mi | 0.07 | 0.3662 |
| 3 | LSA | k09_mx | 0.38 | 0.0380 |
| 3 | LSA | k09_iq | 0.17 | 0.5376 |
| 3 | LSA | k10_md | -0.08 | 0.1888 |
| 3 | LSA | k10_mi | -0.07 | 0.3672 |
| 3 | LSA | k10_mx | 0.18 | 0.4750 |
| 3 | LSA | k10_iq | 0.13 | 0.6614 |
| 3 | LSA | mv05me_md | 0.22 | 0.0525 |
| 3 | LSA | mv05me_mi | 0.13 | 0.2743 |
| 3 | LSA | mv05me_mx | 0.36 | 0.0147 |
| 3 | LSA | mv05me_iq | 0.37 | 0.0044 |
| 3 | LSA | mv10me_md | 0.41 | 0.0033 |
| 3 | LSA | mv10me_mi | 0.22 | 0.0739 |
| 3 | LSA | mv10me_mx | 0.35 | 0.0506 |
| 3 | LSA | mv10me_iq | 0.29 | 0.0625 |
| 3 | LSA | foc_md | 0.11 | 0.9026 |
| 3 | LSA | foc_mi | -0.05 | 0.5935 |
| 3 | LSA | foc_mx | 0.22 | 0.1920 |
| 3 | LSA | foc_iq | 0.17 | 0.6253 |
| 3 | LSA | soc_md | 0.04 | 0.8418 |
| 3 | LSA | soc_mi | 0.15 | 0.5740 |
| 3 | LSA | soc_mx | 0.07 | 0.6198 |
| 3 | LSA | soc_iq | 0.07 | 0.9964 |
| 3 | Word2Vec | k00_md | -0.02 | 0.9076 |
| 3 | Word2Vec | k00_mi | 0.18 | 0.1659 |
| 3 | Word2Vec | k00_mx | 0.02 | 0.8405 |
| 3 | Word2Vec | k00_iq | -0.20 | 0.4775 |
| 3 | Word2Vec | k01_md | 0.19 | 0.0224 |
| 3 | Word2Vec | k01_mi | 0.13 | 0.3960 |
| 3 | Word2Vec | k01_mx | 0.25 | 0.2132 |
| 3 | Word2Vec | k01_iq | 0.10 | 0.3559 |
| 3 | Word2Vec | k02_md | 0.23 | 0.1595 |
| 3 | Word2Vec | k02_mi | 0.13 | 0.3648 |
| 3 | Word2Vec | k02_mx | 0.29 | 0.0862 |
| 3 | Word2Vec | k02_iq | 0.01 | 0.9499 |
| 3 | Word2Vec | k03_md | 0.23 | 0.0832 |
| 3 | Word2Vec | k03_mi | 0.18 | 0.2132 |
| 3 | Word2Vec | k03_mx | 0.23 | 0.2101 |
| 3 | Word2Vec | k03_iq | 0.19 | 0.2039 |
| 3 | Word2Vec | k04_md | 0.22 | 0.1759 |
| 3 | Word2Vec | k04_mi | 0.37 | 0.0046 |
| 3 | Word2Vec | k04_mx | 0.24 | 0.1147 |
| 3 | Word2Vec | k04_iq | 0.14 | 0.3517 |
| 3 | Word2Vec | k05_md | 0.20 | 0.2660 |
| 3 | Word2Vec | k05_mi | 0.12 | 0.3161 |
| 3 | Word2Vec | k05_mx | 0.27 | 0.0504 |
| 3 | Word2Vec | k05_iq | 0.15 | 0.2605 |
| 3 | Word2Vec | k06_md | 0.23 | 0.0554 |
| 3 | Word2Vec | k06_mi | 0.20 | 0.0121 |
| 3 | Word2Vec | k06_mx | 0.01 | 0.6706 |
| 3 | Word2Vec | k06_iq | 0.13 | 0.3396 |
| 3 | Word2Vec | k07_md | 0.22 | 0.1443 |
| 3 | Word2Vec | k07_mi | 0.27 | 0.1004 |
| 3 | Word2Vec | k07_mx | 0.07 | 0.6879 |
| 3 | Word2Vec | k07_iq | 0.04 | 0.6127 |
| 3 | Word2Vec | k08_md | 0.35 | 0.0332 |
| 3 | Word2Vec | k08_mi | 0.38 | 0.0014 |
| 3 | Word2Vec | k08_mx | 0.24 | 0.1856 |
| 3 | Word2Vec | k08_iq | 0.13 | 0.3949 |
| 3 | Word2Vec | k09_md | 0.12 | 0.0916 |
| 3 | Word2Vec | k09_mi | -0.03 | 0.8309 |
| 3 | Word2Vec | k09_mx | 0.25 | 0.0581 |
| 3 | Word2Vec | k09_iq | 0.15 | 0.2172 |
| 3 | Word2Vec | k10_md | 0.09 | 0.0174 |
| 3 | Word2Vec | k10_mi | -0.07 | 0.2294 |
| 3 | Word2Vec | k10_mx | 0.12 | 0.5318 |
| 3 | Word2Vec | k10_iq | 0.07 | 0.5845 |
| 3 | Word2Vec | mv05me_md | 0.30 | 0.0408 |
| 3 | Word2Vec | mv05me_mi | 0.31 | 0.0216 |
| 3 | Word2Vec | mv05me_mx | 0.09 | 0.2171 |
| 3 | Word2Vec | mv05me_iq | -0.14 | 0.8840 |
| 3 | Word2Vec | mv10me_md | 0.42 | 0.0059 |
| 3 | Word2Vec | mv10me_mi | 0.42 | 0.0012 |
| 3 | Word2Vec | mv10me_mx | 0.33 | 0.0306 |
| 3 | Word2Vec | mv10me_iq | 0.07 | 0.8463 |
| 3 | Word2Vec | foc_md | 0.05 | 0.8992 |
| 3 | Word2Vec | foc_mi | -0.01 | 0.8992 |
| 3 | Word2Vec | foc_mx | 0.12 | 0.3413 |
| 3 | Word2Vec | foc_iq | 0.12 | 0.5180 |
| 3 | Word2Vec | soc_md | 0.01 | 0.7905 |
| 3 | Word2Vec | soc_mi | 0.14 | 0.3586 |
| 3 | Word2Vec | soc_mx | 0.09 | 0.6974 |
| 3 | Word2Vec | soc_iq | 0.04 | 0.9128 |
| 3 | RoBERTa | k00_md | 0.24 | 0.0969 |
| 3 | RoBERTa | k00_mi | 0.41 | 0.0038 |
| 3 | RoBERTa | k00_mx | 0.34 | 0.0082 |
| 3 | RoBERTa | k00_iq | -0.14 | 0.3960 |
| 3 | RoBERTa | k01_md | 0.34 | 0.0286 |
| 3 | RoBERTa | k01_mi | 0.48 | 0.0014 |
| 3 | RoBERTa | k01_mx | 0.48 | 0.0013 |
| 3 | RoBERTa | k01_iq | -0.23 | 0.1385 |
| 3 | RoBERTa | k02_md | 0.41 | 0.0046 |
| 3 | RoBERTa | k02_mi | 0.50 | 0.0006 |
| 3 | RoBERTa | k02_mx | 0.63 | 0.0000 |
| 3 | RoBERTa | k02_iq | -0.25 | 0.1030 |
| 3 | RoBERTa | k03_md | 0.34 | 0.0129 |
| 3 | RoBERTa | k03_mi | 0.44 | 0.0028 |
| 3 | RoBERTa | k03_mx | 0.49 | 0.0003 |
| 3 | RoBERTa | k03_iq | -0.25 | 0.1123 |
| 3 | RoBERTa | k04_md | 0.31 | 0.0372 |
| 3 | RoBERTa | k04_mi | 0.45 | 0.0016 |
| 3 | RoBERTa | k04_mx | 0.40 | 0.0011 |
| 3 | RoBERTa | k04_iq | -0.33 | 0.0289 |
| 3 | RoBERTa | k05_md | 0.37 | 0.0046 |
| 3 | RoBERTa | k05_mi | 0.42 | 0.0033 |
| 3 | RoBERTa | k05_mx | 0.45 | 0.0007 |
| 3 | RoBERTa | k05_iq | -0.24 | 0.1223 |
| 3 | RoBERTa | k06_md | 0.30 | 0.0203 |
| 3 | RoBERTa | k06_mi | 0.38 | 0.0109 |
| 3 | RoBERTa | k06_mx | 0.37 | 0.0092 |
| 3 | RoBERTa | k06_iq | -0.33 | 0.0390 |
| 3 | RoBERTa | k07_md | 0.38 | 0.0060 |
| 3 | RoBERTa | k07_mi | 0.43 | 0.0051 |
| 3 | RoBERTa | k07_mx | 0.39 | 0.0006 |
| 3 | RoBERTa | k07_iq | -0.16 | 0.3120 |
| 3 | RoBERTa | k08_md | 0.35 | 0.0148 |
| 3 | RoBERTa | k08_mi | 0.42 | 0.0064 |
| 3 | RoBERTa | k08_mx | 0.41 | 0.0054 |
| 3 | RoBERTa | k08_iq | -0.18 | 0.1728 |
| 3 | RoBERTa | k09_md | 0.29 | 0.0281 |
| 3 | RoBERTa | k09_mi | 0.48 | 0.0006 |
| 3 | RoBERTa | k09_mx | 0.49 | 0.0002 |
| 3 | RoBERTa | k09_iq | -0.18 | 0.2168 |
| 3 | RoBERTa | k10_md | 0.30 | 0.0339 |
| 3 | RoBERTa | k10_mi | 0.39 | 0.0058 |
| 3 | RoBERTa | k10_mx | 0.38 | 0.0021 |
| 3 | RoBERTa | k10_iq | -0.10 | 0.6207 |
| 3 | RoBERTa | mv05me_md | 0.34 | 0.0181 |
| 3 | RoBERTa | mv05me_mi | 0.49 | 0.0011 |
| 3 | RoBERTa | mv05me_mx | 0.39 | 0.0026 |
| 3 | RoBERTa | mv05me_iq | -0.35 | 0.0129 |
| 3 | RoBERTa | mv10me_md | 0.38 | 0.0043 |
| 3 | RoBERTa | mv10me_mi | 0.39 | 0.0081 |
| 3 | RoBERTa | mv10me_mx | 0.34 | 0.0140 |
| 3 | RoBERTa | mv10me_iq | -0.32 | 0.0972 |
| 3 | RoBERTa | foc_md | 0.39 | 0.0585 |
| 3 | RoBERTa | foc_mi | 0.50 | 0.0006 |
| 3 | RoBERTa | foc_mx | 0.27 | 0.2542 |
| 3 | RoBERTa | foc_iq | -0.40 | 0.0117 |
| 3 | RoBERTa | soc_md | 0.30 | 0.1334 |
| 3 | RoBERTa | soc_mi | 0.52 | 0.0002 |
| 3 | RoBERTa | soc_mx | 0.12 | 0.8283 |
| 3 | RoBERTa | soc_iq | -0.33 | 0.0661 |
| 3 | GPT-2 | k00_md | 0.51 | 0.0002 |
| 3 | GPT-2 | k00_mi | 0.63 | 0.0000 |
| 3 | GPT-2 | k00_mx | 0.32 | 0.0080 |
| 3 | GPT-2 | k00_iq | -0.14 | 0.5043 |
| 3 | GPT-2 | k01_md | 0.57 | 0.0000 |
| 3 | GPT-2 | k01_mi | 0.71 | 0.0000 |
| 3 | GPT-2 | k01_mx | 0.08 | 0.4092 |
| 3 | GPT-2 | k01_iq | -0.18 | 0.1380 |
| 3 | GPT-2 | k02_md | 0.80 | 0.0000 |
| 3 | GPT-2 | k02_mi | 0.69 | 0.0000 |
| 3 | GPT-2 | k02_mx | 0.38 | 0.0056 |
| 3 | GPT-2 | k02_iq | 0.03 | 0.8388 |
| 3 | GPT-2 | k03_md | 0.75 | 0.0000 |
| 3 | GPT-2 | k03_mi | 0.77 | 0.0000 |
| 3 | GPT-2 | k03_mx | 0.41 | 0.0026 |
| 3 | GPT-2 | k03_iq | -0.11 | 0.7881 |
| 3 | GPT-2 | k04_md | 0.73 | 0.0000 |
| 3 | GPT-2 | k04_mi | 0.82 | 0.0000 |
| 3 | GPT-2 | k04_mx | 0.37 | 0.0068 |
| 3 | GPT-2 | k04_iq | 0.05 | 0.3884 |
| 3 | GPT-2 | k05_md | 0.78 | 0.0000 |
| 3 | GPT-2 | k05_mi | 0.75 | 0.0000 |
| 3 | GPT-2 | k05_mx | 0.34 | 0.0230 |
| 3 | GPT-2 | k05_iq | 0.21 | 0.1348 |
| 3 | GPT-2 | k06_md | 0.62 | 0.0000 |
| 3 | GPT-2 | k06_mi | 0.72 | 0.0000 |
| 3 | GPT-2 | k06_mx | 0.29 | 0.0241 |
| 3 | GPT-2 | k06_iq | -0.14 | 0.2209 |
| 3 | GPT-2 | k07_md | 0.68 | 0.0000 |
| 3 | GPT-2 | k07_mi | 0.73 | 0.0000 |
| 3 | GPT-2 | k07_mx | 0.26 | 0.0382 |
| 3 | GPT-2 | k07_iq | -0.03 | 0.7023 |
| 3 | GPT-2 | k08_md | 0.60 | 0.0000 |
| 3 | GPT-2 | k08_mi | 0.61 | 0.0000 |
| 3 | GPT-2 | k08_mx | 0.27 | 0.0780 |
| 3 | GPT-2 | k08_iq | 0.12 | 0.7275 |
| 3 | GPT-2 | k09_md | 0.66 | 0.0000 |
| 3 | GPT-2 | k09_mi | 0.62 | 0.0000 |
| 3 | GPT-2 | k09_mx | 0.51 | 0.0003 |
| 3 | GPT-2 | k09_iq | 0.31 | 0.0435 |
| 3 | GPT-2 | k10_md | 0.62 | 0.0000 |
| 3 | GPT-2 | k10_mi | 0.46 | 0.0000 |
| 3 | GPT-2 | k10_mx | 0.46 | 0.0026 |
| 3 | GPT-2 | k10_iq | 0.18 | 0.2226 |
| 3 | GPT-2 | mv05me_md | 0.68 | 0.0000 |
| 3 | GPT-2 | mv05me_mi | 0.67 | 0.0000 |
| 3 | GPT-2 | mv05me_mx | 0.53 | 0.0002 |
| 3 | GPT-2 | mv05me_iq | 0.08 | 0.9513 |
| 3 | GPT-2 | mv10me_md | 0.80 | 0.0000 |
| 3 | GPT-2 | mv10me_mi | 0.78 | 0.0000 |
| 3 | GPT-2 | mv10me_mx | 0.60 | 0.0001 |
| 3 | GPT-2 | mv10me_iq | 0.07 | 0.5066 |
| 3 | GPT-2 | foc_md | 0.52 | 0.0004 |
| 3 | GPT-2 | foc_mi | 0.50 | 0.0015 |
| 3 | GPT-2 | foc_mx | 0.30 | 0.0249 |
| 3 | GPT-2 | foc_iq | -0.20 | 0.1990 |
| 3 | GPT-2 | soc_md | 0.59 | 0.0003 |
| 3 | GPT-2 | soc_mi | 0.66 | 0.0000 |
| 3 | GPT-2 | soc_mx | 0.33 | 0.0211 |
| 3 | GPT-2 | soc_iq | -0.34 | 0.0075 |
| 3 | Llama | k00_md | -0.03 | 0.8874 |
| 3 | Llama | k00_mi | 0.12 | 0.4189 |
| 3 | Llama | k00_mx | 0.11 | 0.1249 |
| 3 | Llama | k00_iq | 0.09 | 0.7389 |
| 3 | Llama | k01_md | 0.17 | 0.2630 |
| 3 | Llama | k01_mi | 0.17 | 0.2836 |
| 3 | Llama | k01_mx | 0.06 | 0.3570 |
| 3 | Llama | k01_iq | 0.12 | 0.3153 |
| 3 | Llama | k02_md | 0.22 | 0.1414 |
| 3 | Llama | k02_mi | 0.13 | 0.3854 |
| 3 | Llama | k02_mx | 0.22 | 0.0445 |
| 3 | Llama | k02_iq | 0.07 | 0.4566 |
| 3 | Llama | k03_md | 0.27 | 0.1187 |
| 3 | Llama | k03_mi | 0.21 | 0.0782 |
| 3 | Llama | k03_mx | 0.21 | 0.1192 |
| 3 | Llama | k03_iq | 0.08 | 0.4401 |
| 3 | Llama | k04_md | 0.22 | 0.1639 |
| 3 | Llama | k04_mi | 0.09 | 0.4414 |
| 3 | Llama | k04_mx | 0.19 | 0.0426 |
| 3 | Llama | k04_iq | 0.17 | 0.2058 |
| 3 | Llama | k05_md | 0.31 | 0.0487 |
| 3 | Llama | k05_mi | 0.15 | 0.2901 |
| 3 | Llama | k05_mx | 0.18 | 0.1995 |
| 3 | Llama | k05_iq | 0.13 | 0.1862 |
| 3 | Llama | k06_md | 0.19 | 0.1876 |
| 3 | Llama | k06_mi | 0.08 | 0.6439 |
| 3 | Llama | k06_mx | 0.15 | 0.3846 |
| 3 | Llama | k06_iq | 0.03 | 0.4769 |
| 3 | Llama | k07_md | 0.20 | 0.1084 |
| 3 | Llama | k07_mi | 0.10 | 0.3954 |
| 3 | Llama | k07_mx | 0.10 | 0.3104 |
| 3 | Llama | k07_iq | 0.04 | 0.3660 |
| 3 | Llama | k08_md | 0.28 | 0.0787 |
| 3 | Llama | k08_mi | 0.06 | 0.8283 |
| 3 | Llama | k08_mx | 0.31 | 0.0333 |
| 3 | Llama | k08_iq | 0.23 | 0.1062 |
| 3 | Llama | k09_md | 0.24 | 0.0650 |
| 3 | Llama | k09_mi | 0.07 | 0.4029 |
| 3 | Llama | k09_mx | 0.34 | 0.0109 |
| 3 | Llama | k09_iq | 0.25 | 0.0438 |
| 3 | Llama | k10_md | 0.29 | 0.0431 |
| 3 | Llama | k10_mi | 0.07 | 0.2738 |
| 3 | Llama | k10_mx | 0.28 | 0.0330 |
| 3 | Llama | k10_iq | 0.28 | 0.1201 |
| 3 | Llama | mv05me_md | 0.13 | 0.3477 |
| 3 | Llama | mv05me_mi | 0.16 | 0.2300 |
| 3 | Llama | mv05me_mx | 0.15 | 0.2267 |
| 3 | Llama | mv05me_iq | 0.06 | 0.9343 |
| 3 | Llama | mv10me_md | 0.26 | 0.1171 |
| 3 | Llama | mv10me_mi | 0.22 | 0.1284 |
| 3 | Llama | mv10me_mx | 0.19 | 0.1890 |
| 3 | Llama | mv10me_iq | 0.05 | 0.7890 |
| 3 | Llama | foc_md | 0.39 | 0.0149 |
| 3 | Llama | foc_mi | 0.46 | 0.0010 |
| 3 | Llama | foc_mx | 0.25 | 0.1065 |
| 3 | Llama | foc_iq | -0.18 | 0.1288 |
| 3 | Llama | soc_md | 0.33 | 0.1111 |
| 3 | Llama | soc_mi | 0.37 | 0.0127 |
| 3 | Llama | soc_mx | 0.13 | 0.5499 |
| 3 | Llama | soc_iq | -0.15 | 0.2418 |
